# Rhinovirus infection of airway epithelial cells uncovers the non-ciliated subset as a likely driver of genetic susceptibility to childhood-onset asthma

**DOI:** 10.1101/2024.02.02.24302068

**Authors:** Sarah Djeddi, Daniela Fernandez-Salinas, George X. Huang, Vitor R. C. Aguiar, Chitrasen Mohanty, Christina Kendziorski, Steven Gazal, Joshua Boyce, Carole Ober, James Gern, Nora Barrett, Maria Gutierrez-Arcelus

## Abstract

Asthma is a complex disease caused by genetic and environmental factors. Epidemiological studies have shown that in children, wheezing during rhinovirus infection (a cause of the common cold) is associated with asthma development during childhood. This has led scientists to hypothesize there could be a causal relationship between rhinovirus infection and asthma or that RV-induced wheezing identifies individuals at increased risk for asthma development. However, not all children who wheeze when they have a cold develop asthma. Genome-wide association studies (GWAS) have identified hundreds of genetic variants contributing to asthma susceptibility, with the vast majority of likely causal variants being non-coding. Integrative analyses with transcriptomic and epigenomic datasets have indicated that T cells drive asthma risk, which has been supported by mouse studies. However, the datasets ascertained in these integrative analyses lack airway epithelial cells. Furthermore, large-scale transcriptomic T cell studies have not identified the regulatory effects of most non-coding risk variants in asthma GWAS, indicating there could be additional cell types harboring these “missing regulatory effects”. Given that airway epithelial cells are the first line of defense against rhinovirus, we hypothesized they could be mediators of genetic susceptibility to asthma. Here we integrate GWAS data with transcriptomic datasets of airway epithelial cells subject to stimuli that could induce activation states relevant to asthma. We demonstrate that epithelial cultures infected with rhinovirus significantly upregulate childhood-onset asthma-associated genes. We show that this upregulation occurs specifically in non-ciliated epithelial cells. This enrichment for genes in asthma risk loci, or ‘asthma heritability enrichment’ is also significant for epithelial genes upregulated with influenza infection, but not with SARS-CoV-2 infection or cytokine activation. Additionally, cells from patients with asthma showed a stronger heritability enrichment compared to cells from healthy individuals. Overall, our results suggest that rhinovirus infection is an environmental factor that interacts with genetic risk factors through non-ciliated airway epithelial cells to drive childhood-onset asthma.

## Introduction

Asthma is a complex and heterogeneous disease that affects 300 million children and adults worldwide and represents a significant burden to healthcare ($82 billion for the US in 2013) ^1^. It is characterized by inflammation of the airways leading to recurrent episodes of airflow obstruction and symptoms such as wheezing, shortness of breath and coughing. Asthma patients have impairments of epithelial barrier function, manifested by irregular disruption of the tight junctions, detachment of ciliated cells and reduced expression of cell-cell adhesion molecules (for instance, E-cadherin) ^2^. This barrier disruption allows environmental substances like allergens, viruses, bacteria and toxic substances to penetrate the submucosa more easily. Allergens, viral infections, and type 2 inflammation have been shown to further damage the barrier integrity of the airway epithelium; moreover, they trigger asthma exacerbations ^3^. Longitudinal epidemiological studies have shown that wheezing caused by rhinovirus infection in children is a risk factor for developing asthma later in childhood ^4–6^. These observations have led to two hypotheses: (1) rhinovirus infection could be causal in asthma development or (2) rhinovirus-induced wheeze is a biomarker that identifies children at increased risk for asthma development. In some children, rhinovirus wheezing does not lead to asthma.

Genome-wide association studies (GWAS) have discovered more than 100 risk loci for asthma ^7,8^. Similar to other complex diseases, the vast majority of the likely causal risk variants are non-coding. As a consequence, deciphering the mechanisms through which the risk alleles lead to disease is challenging; it has been achieved for only a small minority of risk loci. Using a suite of recently developed methods that integrate GWAS data with functional genomics datasets, investigators have discovered key cell types that mediate the genetic susceptibility to complex diseases. For example, risk variants for rheumatoid arthritis are enriched in regulatory elements specific for CD4 T cells, and studies in patients and mice have shown the relevance of these cells in the pathogenesis of this disease ^9–13^. Additionally, GWAS integration with transcriptomics revealed that a significant proportion of the risk alleles for Alzheimer’s disease act through the myeloid lineage rather than the brain ^14^. Alzheimer’s disease is now considered an immune-mediated disease ^14,15^. For asthma, T cell-specific regulatory elements and gene expression are enriched in genetic risk loci ^12,13,16,17^, with some highlighting particularly Th2 cells consistent with the role of type 2 inflammation in asthma pathogenesis ^18–21^.

The observations that non-coding risk variants affect gene regulation in cell types relevant to each disease have motivated large-scale transcriptomic studies to identify genetic variants that are associated with both gene expression (expression quantitative trait loci, eQTL) and disease risk. However, only 25-40% of risk variants for immune-mediated diseases co-localize with eQTLs in immune cells ^22–24^. For asthma, alleles at only 47% of loci co-localize with leukocyte expression and/or splicing QTLs ^22^. Hence, the regulatory effects of most non-coding risk variants remain unknown. More recent studies have highlighted that these “missing regulatory effects’’ could be hidden in specific activation or differentiation cell states that haven’t been systematically ascertained ^25–28^.

GWAS enrichment studies have been highly biased towards annotations of blood immune cell types, with reduced resolution when using other tissues relevant to the context of asthma, such as GTEx tissues from post-mortem human organs ^11,13,16,29,30^. Here we sought to define whether airway epithelial cell states could be driving genetic susceptibility to asthma. We analyzed 10 single-cell and bulk transcriptomic datasets of epithelial cells subject to different activation conditions. We integrated these datasets with GWAS summary statistics for childhood-onset asthma (COA), adult-onset asthma (AOA), unspecified-onset asthma (henceforth referred to as all asthma), and a genetically correlated type 2 inflammation trait: allergy/eczema (**Supplementary Table 1**) ^31–33^. We additionally tested three control traits to assess the specificity of our findings. We used state of the art methods that control for linkage disequilibrium, take advantage of most ascertained genetic variants in the genome, and have been shown to work well for bulk and single cell datasets ^13,17^.

**Supplementary Table 1.** Summary statistics used for heritability enrichment analysis.

|  |  |  |
| --- | --- | --- |
|  | **Studied trait** | **Reference** |
| Asthma-associated traits | Adult-onset asthma | Ferreira et al., Am J Hum Genet, 2019 <sup>38</sup> |
|  | Childhood-onset asthma | Ferreira et al., Am J Hum Genet, 2019 <sup>38</sup> |
|  | Allergy/eczema | UK biobank <sup>65</sup> |
|  | All asthma | UK biobank <sup>65</sup> |
| Control traits | Height | Lango, et al., Nature, 2010 <sup>66</sup> |
|  | Alzheimer's disease | Lambert, et al., Nat Genet, 2013 <sup>67</sup> |
|  | Rheumatoid arthritis | Okada, et al., Nature, 2014<sup>68</sup> |

## Results

We applied two methods that use GWAS data to identify relevant cell types for disease. Linkage Disequilibrium Score-regression in Specifically Expressed Genes (LDSC-SEG) identifies heritability enrichment in genomic annotations (such as genes or chromatin marks with specific presence in a particular cell type or cell state) ^13^. Single-cell disease-relevance score (scDRS) identifies cells, from single-cell RNA-seq data, that significantly express genes in GWAS loci (weighted according to their strength of association with disease) relative to null sets of control genes in the same dataset ^17^. We retrieved GWAS summary statistics from asthma related traits as described above. Throughout our analyses we included three complex traits as controls: height, as a non-immune control, Alzheimer’s disease (AD) as a trait implicating myeloid cells, and rheumatoid arthritis (RA) as a lymphocyte-driven disease with a strong T cell component ^11,13,16,17,29^.

### T cell validation

First, we sought to validate T-cell involvement in the genetic susceptibility to asthma, as previously reported in the literature ^13,16,17,29^. Applying LDSC-SEG to bulk ATAC-seq data of human peripheral blood leukocyte populations, we confirmed that T-cell specific open chromatin regions are significantly enriched in heritability for asthma-related traits (**Supplementary Figure 1**)^16^. Furthermore, when comparing cell types between their resting and activated state, we confirmed that activation-specific open chromatin in T cells has significant heritability enrichment for all the asthma-related traits (**Supplementary Figure 1E**). Next, we applied scDRS to single-cell RNA-seq datasets to identify cells with significant over-expression of risk genes identified from GWAS studies (see Methods). In sinonasal mucosa tissue from healthy donors and chronic rhinosinusitis patients, we observed that 21-83% of cells with significant disease relevant score (10% FDR) for asthma-related traits are T cells (**Supplementary Figure 2**). In a dataset of house dust mite-activated T cells from asthma and allergic patients, we observed that among the cells with significant disease relevant score, most were T effector and Th2 cells (51-57% and 48-42% respectively, **Supplementary Figure 3**). Overall, these analyses confirm the validity of the methods used for this study and confirm previous findings showing the relevance of T cells in the genetic susceptibility to asthma-related traits.

#### Rhinovirus infection induces upregulation of asthma-associated genes in epithelial cells from healthy donors

To assess the role that rhinovirus infection could play in asthma genetic susceptibility at the epithelial cell level, we analyzed a publicly available bulk RNA-seq data of basal airway epithelial cells from healthy donors (N=9) that were infected in vitro with RV-A16 (rhinovirus species RV-A, subtype 16) or treated with PBS vehicle control (**Figure 1A**) ^34^. Using a linear mixed model, we performed differential expression analysis (DEA) testing for rhinovirus infection versus PBS vehicle. From the 14,883 tested genes, we selected the top 10% based on t-statistic (as recommended by LDSC-SEG) to select genes that are upregulated with rhinovirus infection(**Figure 1B**). We used LDSC-SEG to investigate whether this gene set showed an enrichment of asthma heritability. Our analysis showed significant heritability enrichment in rhinovirus-upregulated genes for all asthma (P = 0.033) and COA (P = 0.037), with a larger coefficient observed for COA (**Figure 1C**). Moreover, we did not observe any significant enrichment for any of the control traits (AD, RA, height) (**Supplementary Figure 4A**). The fact that we did not observe significant heritability enrichment in RV-upregulated genes for RA suggests that the signal observed for all asthma and COA is not due to a general immune transcriptional response, but rather a response that is specific for RV-infection of epithelial cells.

**Figure 1.**
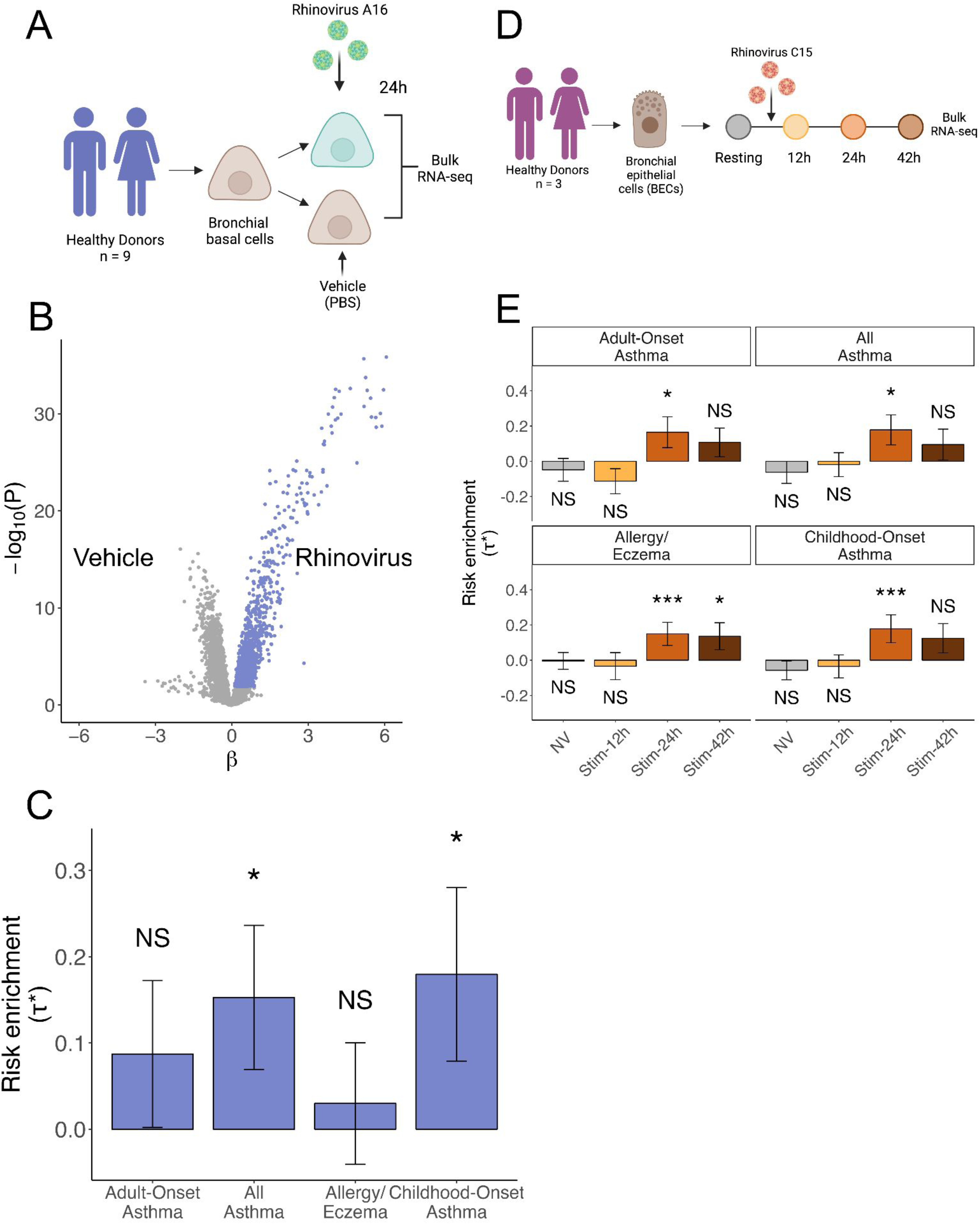
Bronchial epithelial cells infected with rhinovirus upregulate genes associated with asthma susceptibility. **(A)** Experimental design of the *Helling et al.* dataset consisting of basal bronchial epithelial cells from healthy donors stimulated with PBS or RV-A16. **(B)** Volcano plot showing differentially expressed genes after rhinovirus infection, genes selected based on t-statistic are colored in purple. **(C)** Bar plot showing LDSC-SEG heritability enrichment coefficient (τ*) for each of the asthma-associated GWAS studies. Error bars represent τ* +/-standard error. Asterisk denotes P < 0.05 and NS denotes nonsignificant (P > 0.05). **(D)** Experimental design of the Basnet et al. time course dataset consisting of BECs stimulated with RV-C15. **(E)** Bar plot showing LDSC-SEG heritability enrichment coefficient (t*) for differentially expressed genes at each time point when compared against all others. Error bars represent τ* +/-standard error. Asterisks denote significance as * P < 0.05, *** Bonferroni-adjusted P < 0.05 and NS denotes nonsignificant (P > 0.05).

Next, we sought to validate these findings in an independent study using a different strain of RV. We reanalyzed a bulk RNA-seq time course dataset of epithelial cells from 3 healthy donors where cells were infected with RV-C15, and samples were collected before infection and at 12, 24, and 42 hours post-infection (HPI) (**Figure 1D**) ^35^. We performed differential expression analysis to identify genes upregulated specifically in each time point (versus all others), and applied LDSC-SEG (**Supplementary Figure 4B**). We observed significant heritability enrichment for upregulated genes by rhinovirus infection specifically at 24 hours for all asthma-related traits (P < 0.05, **Figure 1E**). This enrichment was higher for COA and allergy/eczema, compared to all asthma and AOA (**Figure 1E**). Genes that were specifically expressed at 42 hours post-infection also had positive enrichment for heritability in all traits, but this was only significant for allergy/eczema (P = 0.03). Once again, this enrichment was not present in any of the control traits (**Supplementary Figure 4C**). Together, these findings suggest that rhinovirus-infected epithelial cells represent a cell state that may mediate genetic susceptibility to asthma, with a greater contribution to COA than to AOA.

#### Asthma-associated genes after rhinovirus infection are specifically enriched in non-ciliated epithelial cells

We then asked whether there were specific epithelial cell subsets that may mediate asthma genetic risk after rhinovirus infection. To evaluate this, we used single-cell RNA-seq data of 3 healthy donors, where airway epithelial cell samples were infected with RV-C15 or resting and profiled at 24 hours (**Figure 2A**, **Supplementary Figure 5A-C**). We performed cell clustering and then annotated the clusters based on epithelial cell markers (**Supplementary Figure 5D**). We identified 2 ciliated cell subsets, and 5 non-ciliated cell subsets: basal, deuterosomal, neuroendocrine, secretory, and transitional (**Figure 2B**, **Supplementary Figure 5D**). We applied scDRS on this dataset to identify cells with significant over-expression of asthma-associated genes. The number of cells with significant disease relevant scores (10% FDR) varied per trait: 147 cells for all asthma, 850 cells for COA, 0 for AOA and 147 cells for allergy/eczema (**Figure 2C**). Notably, COA was the trait with the highest proportion of disease-relevant cells. Among the cells with a significant disease relevant score, 99% corresponded to the stimulated condition. Furthermore, the disease relevant cells were strongly over-represented in the non-ciliated cell subsets, representing 96-99% of the significant cells, while they make up 25% of the whole dataset (**Figure 2C-D**). As expected, we did not observe any significant cells for height, AD and RA (**Supplementary Figure 5E**).

**Figure 2.**
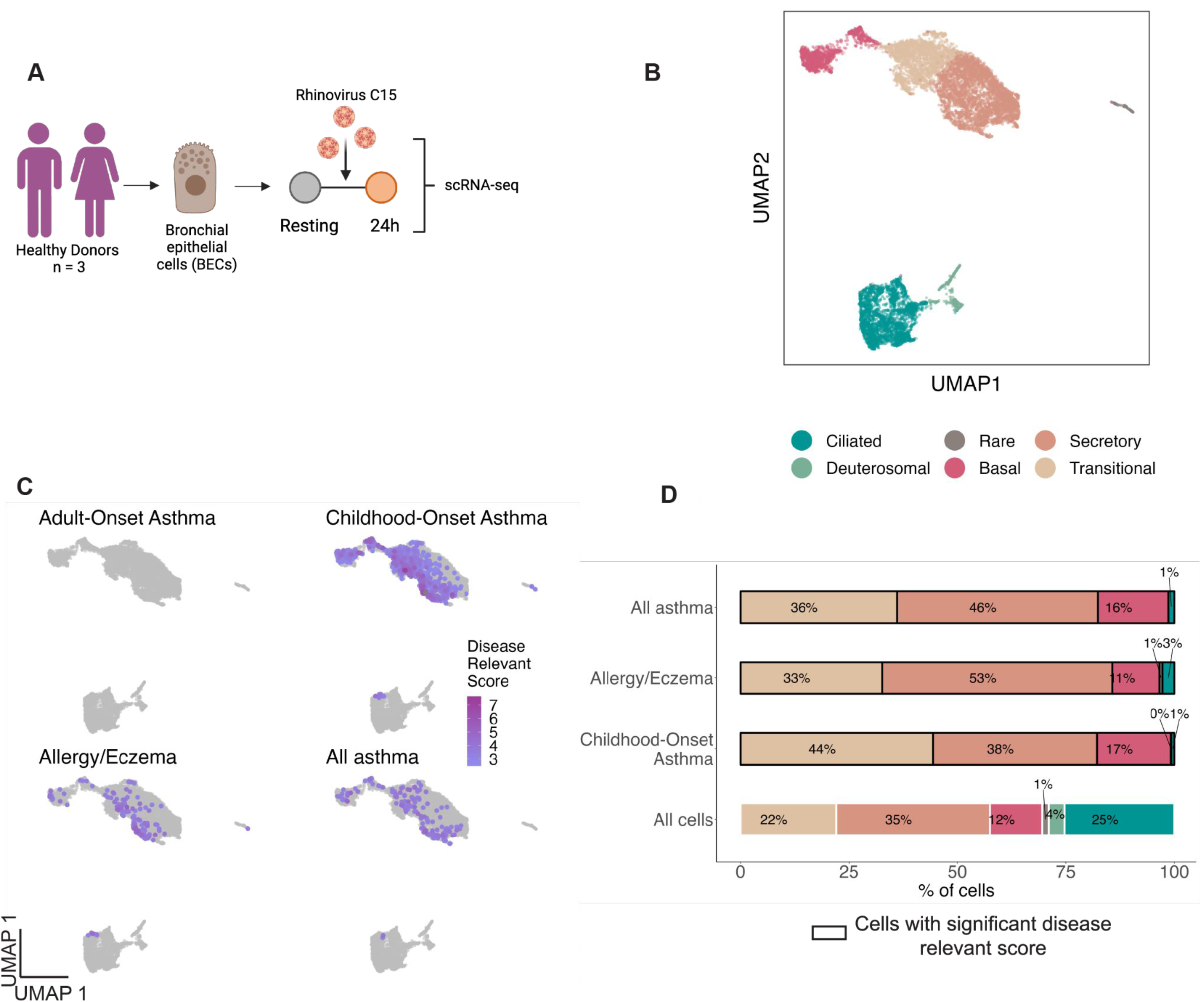
scRNA-seq uncovers non-ciliated epithelial cells as potential mediators for asthma risk. **(A)** Experimental design of the *Basnet et al.* scRNA-seq dataset of BECs from healthy donors infected with RV-C15. **(B)** UMAP visualization of the 10,721 airway epithelial cells colored by cell type. **(C)** scDRS results represented on the UMAP for the 4 asthma-associated GWAS tested. The intensity of the color represents the disease relevant score, the lighter purple represents a less intense score whereas a more intense purple represents cells associated with a stronger score. nonsignificant cells with a FDR higher than 10% are depicted in gray. **(D)** Bar plot representing the percentage of each cell type in all cells followed by the significant cells at 10% FDR for scDRS in COA, Allergy/Eczema, All asthma. AOA is not represented on this barplot because no cells were significant.

Overall, we identified non-ciliated cells as the main epithelial cell subset with significant upregulation of childhood-onset asthma-associated genes after rhinovirus infection. However, only ciliated cells are known to be infected by RV-C15, which we confirmed by looking at the expression of the RV-15 receptor (*CDHR3*) and the presence of the viral sequence itself in the scRNA-seq data ^36^ (**Supplementary Figure 5F-G**). We therefore hypothesized that ciliated cells may communicate with non-ciliated cells upon rhinovirus infection. We used CellphoneDB to investigate some of the possible ligand-receptor mechanisms through which cells may be communicating ^37^. Specifically, we looked for ligand-encoding genes expressed in ciliated cells and their corresponding receptor-encoding gene expressed in non-ciliated cells. Furthermore, we required that the ciliated cell ligand-encoding gene is upregulated upon rhinovirus infection. We identified a potential pair of interactors consisting of *LGALS9*, which codes for a galectin from the beta-galactoside-binding protein family implicated in the modulation of the cell-cell and cell-matrix interactions, and *SORL1*, a gene encoding sortilin-related receptor, which may have a role in endocytosis and intracellular trafficking (**Supplementary Figure 5H-I**).

#### Enrichment of asthma-associated genes after rhinovirus infection is strong in epithelial cells from asthma patients

Having observed the enrichment of asthma-associated genes after RV infection in both bulk and single-cell level in healthy subjects, we asked whether we would observe the same enrichment in samples coming from asthma patients. To do this, we repeated the differential expression analysis between rhinovirus RV-A16 infection and treatment with PBS from the first dataset ^34^, this time using the asthma patient cohort (**Figure 3A**). We found 2,843 differentially expressed genes at 5% FDR, 1,353 of them upregulated and 1,481 downregulated after RV infection (**Figure 3B**). Of the 2,843 differentially expressed genes in patients, 1,834 were also differentially expressed in healthy controls. After selecting the top 10% genes by t-statistic (1,488) and running LDSC-SEG, we observed significant heritability enrichment for all asthma (P = 0.003) and COA (P = 0.003, **Figure 3C**). We did not observe heritability enrichment for down-regulated genes by RV (P > 0.05, **Supplementary Figure 6C**). While the results were consistent with what we observed in healthy controls (COA τ*=0.17, all asthma τ*= 0.15), the enrichment of RV-upregulated asthma-associated genes was more significant and had a larger enrichment coefficient in the asthma patients (COA τ*=0.32, all asthma τ*=0.25).

**Figure 3.**
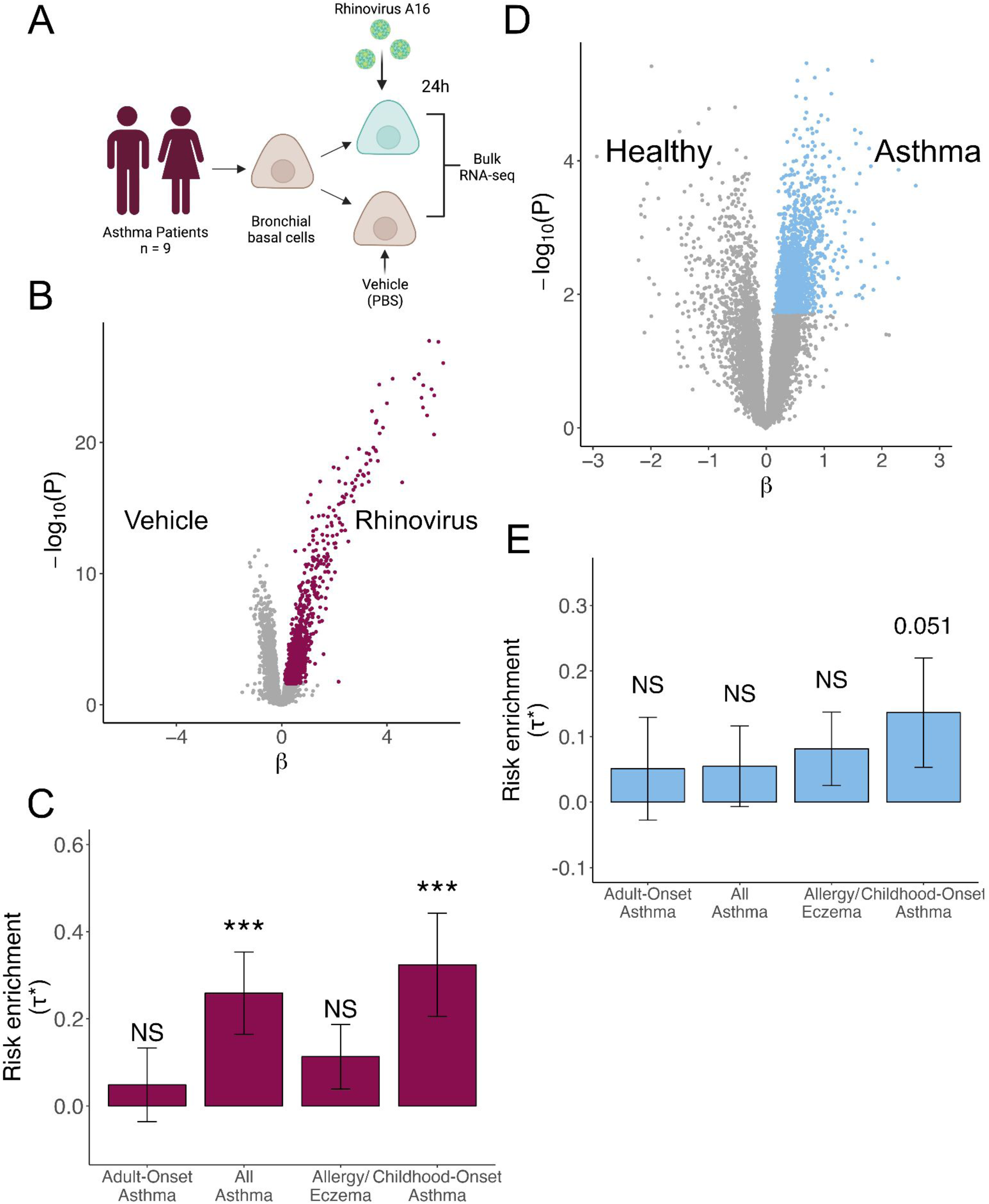
RV-infected BECs from asthma patients showed a stronger enrichment for asthma risk compared to those of healthy individuals. **(A)** Experimental design of *Helling et al.* dataset consisting of BECs from asthma patients stimulated with PBS or RV-A16. **(B)** Volcano plot showing differentially expressed genes after rhinovirus infection in patient samples. Genes upregulated after infection were selected based on t-statistic and are colored in burgundy. **(C)** Bar plot showing LDSC-SEG heritability enrichment coefficient (τ*) across GWAS studies. Error bars represent τ* +/-standard error. Asterisks denote significance as *** Bonferroni-adjusted P < 0.05 and NS denotes nonsignificant (P > 0.05). **(D)** Volcano plot showing differentially expressed genes between asthma patients and healthy donors. Genes upregulated in patients were selected based on t-statistic and are colored in blue. **(E)** Bar plot of LDSC-SEG heritability enrichment coefficient (τ*), suggestively significant enrichment is labeled. Error bars represent τ* +/-standard error. NS denotes nonsignificant (P > 0.05).

Based on these results, we hypothesized that asthma patients might have airway epithelial cells in a transcriptomic state that over-expresses asthma-risk genes in comparison to healthy controls, which might be linked to or independent of their response to RV. To test this, we analyzed differentially expressed genes between asthma patients and healthy individuals taking all samples while controlling for RV/PBS treatment. We found 994 differentially expressed genes between patients and controls (5% FDR, **Figure 3D**). After selecting the top 10% genes upregulated in patients based on t-statistic (1,593), we found a suggestive significant enrichment for COA heritability (P = 0.05, **Figure 3E**). As expected, our control traits did not have any significant heritability enrichment for either of the annotations tested (**Supplementary Figure 6**). Overall, these results suggest that the epithelial cells from patients could be in a state that is over-expressing asthma-associated genes.

#### Genes at asthma risk loci upregulated with rhinovirus infection in airway epithelial cells

We then investigated which of the genes upregulated by rhinovirus infection are associated with COA and AOA. To do so, we retrieved the GWAS lead variants identified by Ferreira et al. ^38^. We then linked risk variants to genes using three approaches: (1) selecting the likely target genes identified by the locus-to-gene (L2G) algorithm of Open Targets Genetics, herein called L2G genes, (2) selecting the closest gene to the lead variant, and (3) selecting genes within a 250kb window of the lead variant (see Methods). From these gene lists, we selected genes that were upregulated upon rhinovirus infection in epithelial cells at 5% FDR (**Figure 4**) ^34,35^. For COA we identified 55 risk loci with genes upregulated upon rhinovirus infection in epithelial cells, 13 of which have L2G likely target genes (e.g. *IL1RL1*, *IL4R*, *GSDMB*, *OVOL1*, *MYC*), and 6 are the closest gene to the lead variant (e.g. *IRF1*, *GPR183*). For AOA only 19 risk loci have genes upregulated by RV in epithelial cells, among which 3 are likely target genes (e.g. *IL4R*, *HDAC7*, *IL1RL1*) and 3 are the closest gene to the lead variant (e.g. *RAPGEF3*, *IRF1*, *SSR3*). Few genes were shared between COA and AOA (*IRF1*, *IL4R*, *PDLIM4*, *IL1R2* and *IL1RL1*).

**Figure 4.**
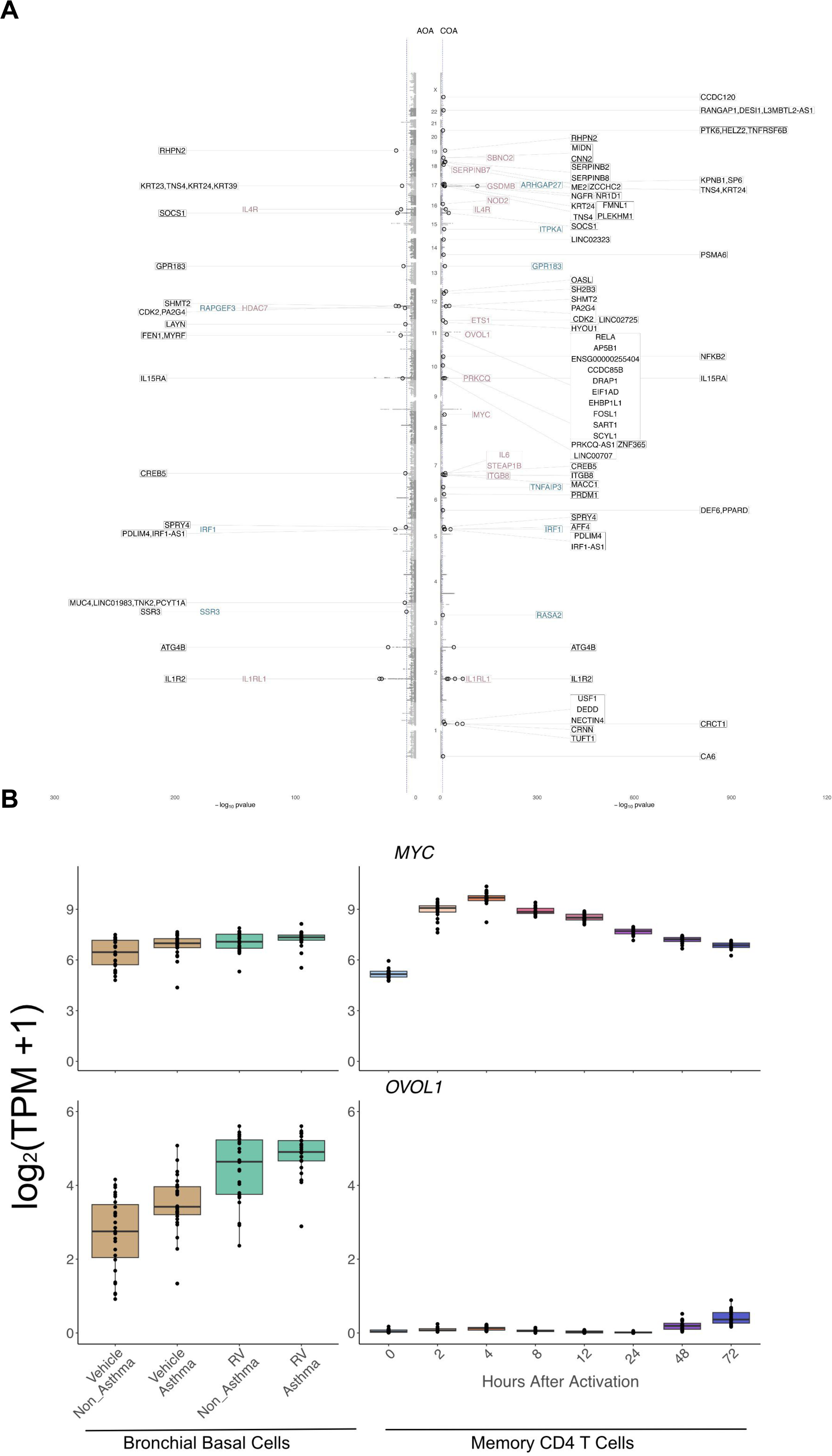
Rhinovirus induced genes that are found in COA and AOA GWAS. **(A)** Miami plot of COA and AOA GWAS. Each gray dot shows the SNP found in the *Ferreira et al.*, GWAS. The black circles represent SNPs being found as lead variants in either Open Targets Genetics or in Ferreira study. Highlighted genes are upregulated upon rhinovirus infection, in purple the genes being L2G genes, in blue the closest one to the transcription start site of the variant (Open Target Genetics), and in black genes found in a window of 250kb around the SNP. The blue dashed line represents the P-value threshold of −log10(5×10^−8^). **(B)** Box plots depicting gene expression levels for *MYC* (top) and *OVOL1* (bottom). Left panel shows gene expression in epithelial cells from the *Helling et al.* dataset; samples infected with RV-A16 are shown in green and non-infected samples are in brown. Right panel shows gene expression from *Gutierrez-Arcelus et al.* in activated CD4 memory T cells; colors depict time points after activation. In each plot every point represents an individual sample.

After having characterized the asthma-associated genes that are upregulated in RV-infected epithelial cells, we sought to define which of these genes are shared with T cells. To do so, we compared the levels of expression of the genes in the RV-datasets with a dataset we previously published consisting of 8 activation time points of human periphery memory CD4+ T cells stimulated with anti-CD3/CD28 microbeads ^25^. One of the highlighted genes that also had an increased expression in T cells was *MYC*, with an increase at 2 and 4 hours post-stimulation. Notably, this gene’s expression was not only increased after rhinovirus infection within asthma patients (P=0.01) and within healthy controls (P=0.0009) but was also upregulated in patients compared to controls (P=0.01). On the other hand, *OVOL1*, another GWAS gene upregulated in RV-infected epithelial cells, shows the same pattern of expression as *MYC* in epithelial cells, but shows almost no expression in T cells (**Figure 4B**, **Supplementary Figure 7**). *OVOL1* is also associated with atopic dermatitis, another type 2 inflammatory disease and a recent meta-analysis study confirmed this susceptibility locus for eczema-associated asthma ^39^.

#### Other viral infections in epithelial cells and their association with asthma susceptibility

We next asked whether other viruses could potentially be inducing upregulation of asthma-associated genes in epithelial cells. First, we analyzed a bulk RNA-seq dataset of bronchial epithelial cells stimulated by influenza A virus at 48 hours or sham control (N = 3 healthy donors, **Figure 5A**). We identified differentially expressed genes between influenza A and the sham control and we selected the top 10% upregulated genes ranked by a t-statistic to run LDSC-SEG (**Supplementary Figure 8A**). The results show a significant enrichment of heritability for the four asthma-related traits (P < 0.05), suggesting that influenza A infection also significantly upregulates asthma and allergy-associated genes (**Figure 5B**). We did not observe any significant enrichment for the control traits (**Supplementary Figure 8B**).

**Figure 5.**
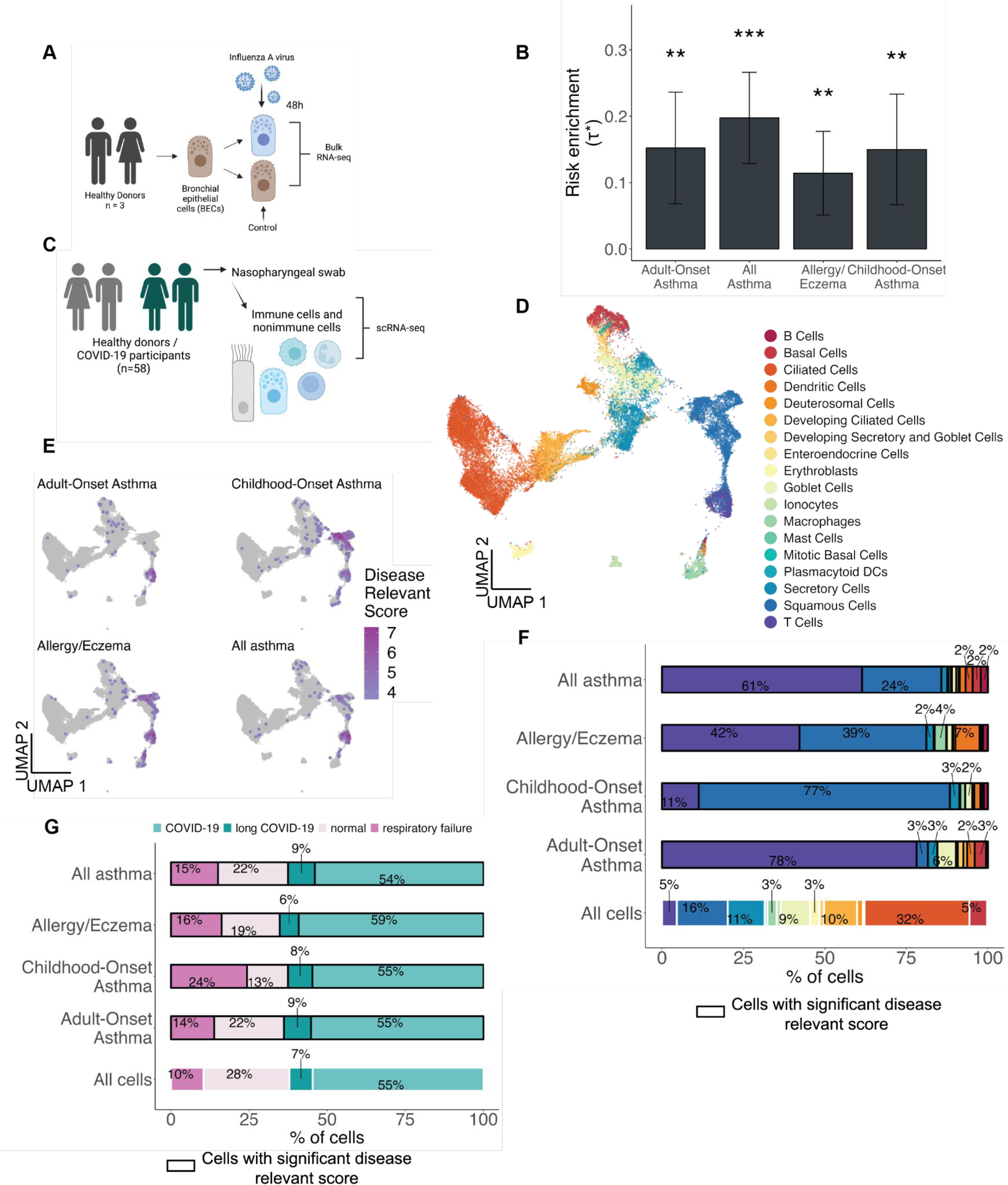
Influence from other viruses on asthma genetics. **(A)** Experimental design of *Tao et al.* bulk RNA-seq dataset. **(B)** Bar plot representing LDSC-SEG enrichment. Error bars represent τ* +/-standard error. Asterisk denotes significance as * P < 0.05. **(C)** Experimental design of *Ziegler et al.* single-cell RNA-seq dataset of epithelial cells infected with SARS-CoV-2 or not. **(D)** UMAP visualization of the 32,588 cells colored by cell type. **(E)** scDRS results represented on the UMAP for the 4 asthma-associated GWAS tested. The intensity of the color represents the disease relevant score, the lighter purple represents a less intense score whereas a more intense purple represents cells associated with a stronger score. nonsignificant cells with a FDR higher than 10% are depicted in gray. **(F)** Bar plot representing the percentage of each cell type in all cells followed by the significant cells at 10% FDR for scDRS in AOA, COA, Allergy/Eczema and All asthma. **(G)** Bar plot representing the percentage of cells in the full dataset coming from patients grouped by COVID severity categories (COVID-19, long COVID-19, respiratory failure) or from healthy donors (normal). Followed by percentage of cells passing significance at 10% FDR for AOA, COA, Allergy/Eczema, All asthma, by disease severity category or control.

Subsequently, we analyzed a single-cell RNA-seq dataset of immune and non-immune cells obtained by nasopharyngeal swabs from COVID-19 patients or healthy donors (N=58, **Figure 5C-D**) ^40^. We identified cells with significant disease-relevant scores at 10% FDR, among which we found 174 cells for AOA, 795 cells for COA, 594 for allergy/eczema and 280 for all asthma (**Figure 5E**). COA yielded the highest number of disease relevant cells. We observed an increase in proportions of enriched cells for T cells in all traits and for squamous cells in COA, allergy/eczema and all asthma (**Figure 5F**). More precisely, T cells which represented 5% of the cells in the dataset, constituted 78% of significant cells for AOA, 11% for COA, 42% for allergy/eczema and 61% for all asthma (**Figure 5F**). The COVID-19 status did not have any significant impact on the asthma-associated expression, as the disease-relevant cells were not overrepresented in any specific patient group (**Figure 5G**). As expected, we observed an enrichment for the macrophages cluster in Alzheimer’s and T cells cluster in rheumatoid arthritis (**Supplementary Figure 8C**). Additionally, we analyzed a dataset of bronchial epithelial cells (BECs) infected with SARS-CoV-2 in vitro, and profiled with scRNA-seq after one, two or three days, along with non-infected cells (N = 1 healthy control, **Supplementary Figure 9A-B**). We clustered and annotated ciliated and non-ciliated cell subsets, and applied scDRS for the four asthma-related traits (**Supplementary Figure 9C-D**). We identified cells with significant disease-relevant scores at 10% FDR, finding 3 cells for AOA, 802 cells for COA, 243 cells for allergy/eczema and 621 cells for all asthma (**Supplementary Figure 9D-E**). The disease relevant cells were strongly over-represented in the non-infected cells subsets, constituting 62-89% of the significant cells (**Supplementary Figure 9F**). Overall, these findings indicate that the influenza virus significantly induces the expression of asthma-associated genes, whereas SARS-CoV-2 does not.

### Epithelial cells activated with cytokines relevant for type 2 inflammation

Both epithelial and immune cells respond to cytokines by upregulating signaling pathways that drive inflammation. Some pro-inflammatory cytokines relevant to asthma are IL-4, IL-13, IL-17 and interferon (IFNγ). These cytokines are upregulated in subsets of patients with severe or type 2 asthma ^41–46^. Moreover, blockade of IL4Rα is a highly effective treatment for moderate to severe asthma ^47^. Consequently, we asked whether epithelial cells stimulated with cytokines might induce a transcriptional program enriched for asthma-associated genes. First we used a bulk RNA-seq dataset consisting of human bronchial epithelial cells (HBECs) that were stimulated with either IFNα, IFNγ, IL-13 or IL-17 (N = 6 healthy donors, **Figure 6A**) ^48^. We selected the top 10% upregulated genes by t-statistic for each stimulus (**Supplementary Figure 10A**). We used LDSC-SEG to analyze these 4 sets of genes in the 4 asthma-associated traits and the control traits. We did not identify any significant heritability enrichment for any of the stimuli gene sets for the asthma-associated traits tested here (**Figure 6B**), nor for the control traits (except for genes upregulated by IFNγ for rheumatoid arthritis, **Supplementary Figure 10B**).

**Figure 6.**
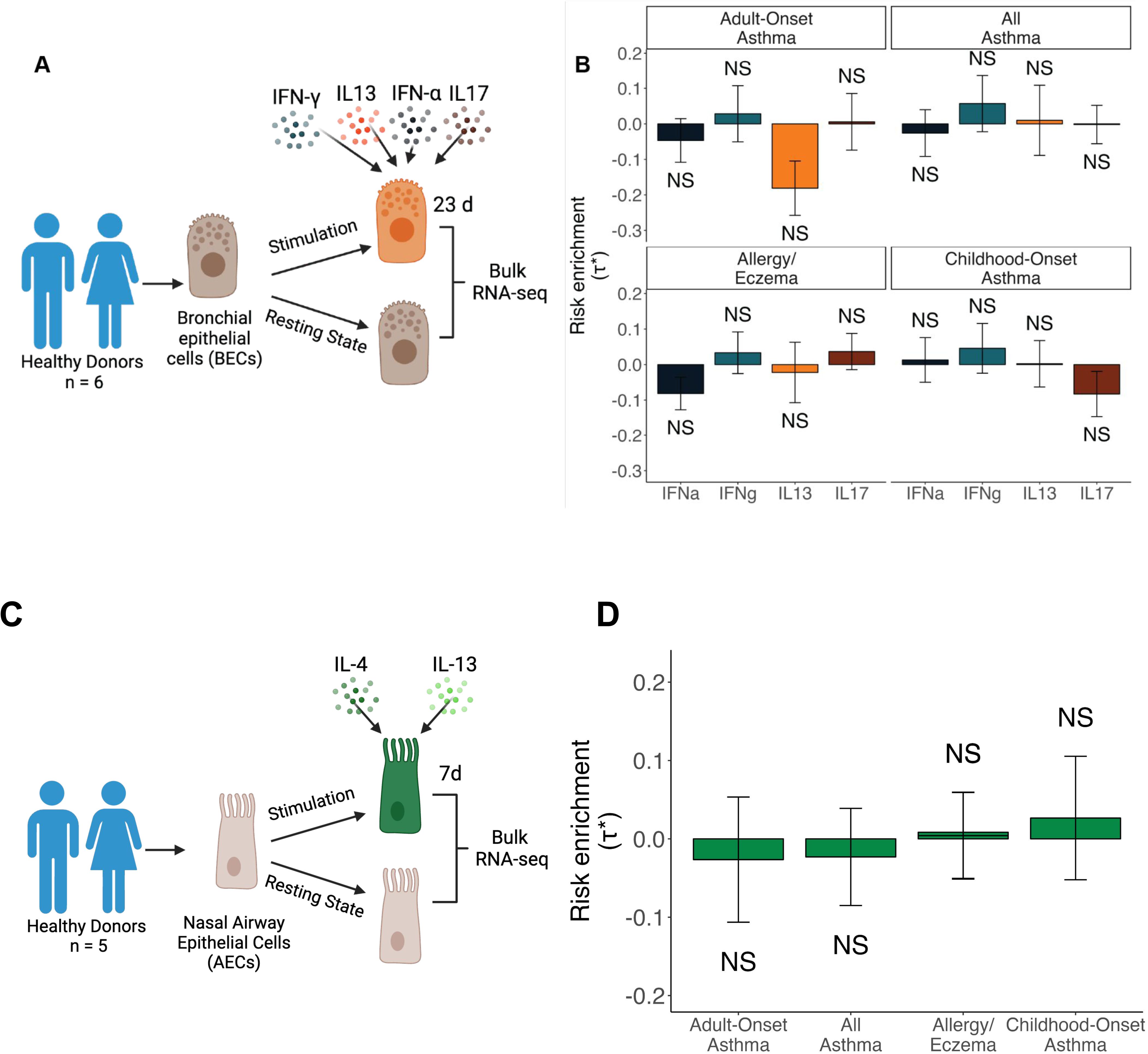
Cytokines impact on asthma-associated genes. **(A)** Experimental design of the *Koh et al.* bulk RNA-seq dataset of BECs from healthy donors that were stimulated or not with different cytokines (IFNγ, IFNɑ, IL-13, IL-17). **(B)** Bar plot representing LDSC-SEG heritability enrichment coefficient (τ*) for genes up-regulated in each stimuli against all others, for each of the asthma-associated GWAS.Error bars represent τ* +/-standard error. NS denotes nonsignificant (P > 0.05). **(C)** Experimental design of bulk RNA-seq of AECs from healthy donors co-stimulated with IL-4 and IL-13. **(D)** Bar plot showing LDSC-SEG enrichment for each of the asthma-associated GWAS. Error bars represent τ* +/-standard error. NS denotes nonsignificant (P > 0.05)

Next, given that IL-13 might work synergistically with IL-4, we performed bulk RNA-seq of nasal airway epithelial cells from healthy donors (N = 5) co-stimulated in vitro with IL-4 and IL-13 (**Figure 6C**). We tested DE genes for the IL-4-IL-13 condition compared to the unstimulated control. In line with the results observed in the previous analysis, we found no significant enrichment for any of the asthma-associated (**Figure 6D**) or control traits (except for AD, **Supplementary Figure 10E**). Together, these results suggest that epithelial cells upregulate asthma-associated genes in a stimulus-specific manner, which to the extent of this study, is not caused by the stimulation with the cytokines tested here.

## Discussion

While some genetic risk variants for asthma are enriched near genes with T cell-specific expression ^13,17,22,28^, the effects of most variants on gene regulation remain unknown. In this study, we asked whether some of these “missing regulatory effects” could be hidden in airway epithelial cells, given they are the first line of contact for respiratory viruses, including those that have been associated with asthma development or exacerbations. We analyzed ten transcriptomic datasets of human airway epithelial cells cultured under different stimuli and integrated them with genetic susceptibility data for asthma and related traits. We consistently showed that rhinovirus-activated epithelial cells significantly upregulate genes at childhood-onset asthma risk loci. We observed this in samples from healthy donors and even more so in cells from asthma patients. Notably, we discovered that non-ciliated cells are the subset driving these associations with asthma, indicating that non-ciliated airway epithelial cells activated with rhinovirus are key mediators of genetic susceptibility to childhood-onset asthma. While other respiratory viruses, such as influenza might also significantly upregulate genes at asthma risk loci, this is not likely a general virus response or epithelial cell activation signature, given that we did not detect asthma heritability enrichment for SARS-CoV-2 or cytokine-upregulated genes.

Our findings are consistent with epidemiological studies that have shown associations between wheezing illness caused by rhinovirus infection and asthma development in children ^4,49–51^. Additionally, a previous birth cohort study identified genetic variants at the 17q21 locus that were associated with asthma in children who had rhinovirus-associated wheezing illness in the first 3 years of life, but not in children who had RSV-associated wheezing illnesses at those same ages ^52^. In that study, rhinovirus upregulated two genes at this locus, *ORMDL3* and *GSDMB*, in PBMCs ^52^. Here, we observe that in non-ciliated airway epithelial cells rhinovirus induces upregulation of *GSDMB* as well as putative causal genes in 54 additional loci. This demonstrates a widespread interaction between *in vitro* rhinovirus infection and polygenic susceptibility to childhood-onset asthma, specifically mediated through airway epithelial cells. These findings are concordant with a previous study reporting that genes at COA-specific risk loci (as compared to AOA) have high expression in skin, which is a barrier tissue with an abundance of epithelial cells^30^. Overall, our findings support the hypothesis that rhinovirus could be causally linked to asthma development in children and not just be a biomarker of children destined to develop asthma. Not all children that get RV-wheezing develop asthma, and our findings suggest that the combination of preschool rhinovirus wheezing illnesses and a high genetic burden synergistically promote the development of childhood asthma.

We discovered that non-ciliated cells (basal, secretory, and transitional) are the specific cell subsets that overexpress genes at asthma risk loci. This suggests that an important fraction of the non-coding risk variants for asthma likely affect gene regulation in non-ciliated cells under specific viral activation states. In our study we looked at two different rhinovirus types. For the case of RV-C15, the receptor of the virus, CDHR3, is mainly expressed in the ciliated cells ^53,54^ and viral RNA quantification confirmed this subset is the one directly infected by the virus (**Supplementary Figure 5G**). This suggested that RV-infected ciliated cells efficiently transmit a signal to the non-ciliated cells, which then express genes in asthma risk loci. The time course experiments indicated this upregulation of asthma-associated genes occurred predominantly at 24 and 42 hours ^35^. In our analyses of RV-A16 infected epithelial cells, the data came from bulk RNA-seq of basal cells (non-ciliated) treated for 24 hours. In contrast to RV-C15, RV-A16 binds to the ICAM receptor, which is expressed in ciliated cells and basal cells ^55^. Strikingly, while the cell subsets that get directly infected differ between the two RV strains, both significantly upregulated asthma-associated genes in non-ciliated cells at 24 hours. By contrast, for SARS-CoV-2, *in vitro* infection did not upregulate genes in asthma risk loci; rather, the non-infected cells presented a significant expression of asthma-associated genes. Although the data in this study came from only one individual, the cells with significant disease relevant scores were also predominantly non-ciliated cells (> 83%, **Supplementary Fig. 9**). Future single-cell studies with larger sample sizes and ascertaining infection by multiple types of viruses could point to additional epithelial cell subsets and cell states as candidate drivers of genetic susceptibility to asthma.

The observations in our study may also be relevant to virus-induced asthma exacerbation ^56^. Here, we not only demonstrate that rhinovirus infection induces a transcriptional response enriched in childhood-onset asthma risk, but we also identified a heritability enrichment for genes upregulated in asthma patients compared to controls, even when controlling for RV infection (suggestive P = 0.051, **Figure 3E**). This result goes in line with a previous observation that, at the open chromatin level, airway epithelial cells of asthma patients have a large amount of open chromatin regions at baseline that are RV-response regions in healthy controls ^34^. It is possible that over-expression of asthma-associated genes at baseline may increase the risk for acute virus-induced exacerbations in patients with asthma. Influenza infections, which can cause asthma exacerbations (especially in adults) ^57–59^, induced an enrichment of both adult-onset and childhood-onset asthma heritability in influenza upregulated genes in airway epithelial cells. Furthermore, SARS-CoV-2 seems less likely than other viruses to provoke asthma exacerbations, and asthma does not appear to be a risk factor for severe SARS-CoV-2 infection ^60^. This could be due to multiple reasons, such as allergy-induced reduction in the ACE2 receptor ^61^, but it is also in line with our observations that SARS-CoV-2 infection itself does not induce a transcriptional program significantly enriched in asthma heritability ^61^.

Our study had some limitations. We were not able to ascertain all possible epithelial cell subsets and states. Most of the datasets analyzed involved *in vitro* infections, rather than *in vivo* infected samples. Additionally, all samples came from adults, which made it all the more striking that we detected heritability enrichments for childhood-onset asthma. Future studies in children are important to validate these findings. Furthermore, we were limited by the cell sources and specific time points and experimental designs of each study. In particular, the absence of asthma heritability enrichment in cytokine-upregulated genes in epithelial cells could imply multiple scenarios. One possibility could be that even though pro-inflammatory cytokines (IL-4, IL-13, IFNα, IFNγ, IL-17) upregulate many genes in epithelial cells (684-2876 at 5% FDR in our analyses), they do not significantly interact with polygenic risk factors for asthma in epithelial cells. Other possibilities for the absence of signal could be that the cytokine-induce activation might interact with genetic risk factors acting in T cells or other non-epithelial cells, or that there are interactions with environmental conditions not present in the models included in this analysis.

Overall, our findings of asthma heritability enrichment in various epithelial cell states (resting versus virus-infected, patients versus healthy controls) could reflect variability in how risk variants contribute to disease onset versus progression. These results highlight the importance of studying the cellular context in which GWAS loci contribute to disease risk and will ultimately help to better understand the mechanisms through which those risk variants are acting. Moreover, the outcomes of our study could open the door to new therapeutic avenues. Indeed, drug targets that have genetic evidence are more likely to be approved and move forward to clinical trials than those without it ^62^. Large-scale multi-omic studies (with comparable power to GWAS ^63^) of non-ciliated airway epithelial cells activated with rhinovirus could help identify the target genes of non-coding asthma risk variants, together with functional validations with approaches such as base editing. Finally, if our observations are confirmed and further characterized by future studies, it would support the development of a rhinovirus vaccine or other protective intervention as a way to prevent childhood-onset asthma ^64^.

## Material and methods

### Dataset collection

We downloaded transcriptomic datasets from the National Center for Biotechnology Information (NCBI) Gene Expression Omnibus (GEO), Genome Sequence Archive (GSA) and from ImmPORT. We also downloaded a chromatin accessibility dataset (ATAC-seq) from GEO (**Supplementary Table 2**).

**Supplementary Table 2.** Datasets used in this study.

| **Supplementary Table 2. Datasets used in this study.** |  |  |
| --- | --- | --- |
| **Data type** | **Source** | **Accession number** |
| ATAC-seq | Calderon et al., 2019 | GSE118189 |
| scRNA-seq | Wang et al., 2023 | HRA000772 (Genome Sequence Archive) |
| scRNA-seq | Seumois et al., 2020 | GSE146170 |
| RNA-seq | Helling et al., 2020 | GSE152550 |
| RNA-seq | Basnet et al., 2023 | SDY1882 (ImmPORT) |
| scRNA-seq | Basnet et al., 2023 | SDY1882 (ImmPORT) |
| RNA-seq | Tao et al., 2022 | GSE193164 |
| scRNA-seq | Ravindra et al., 2021 | GSE166766 |
| RNA-seq | Koh et al., 2022 | GSE185200 |
| RNA-seq | Barret lab | Will be published as part of current study |

### Ethical approval

The Mass General Brigham Institutional Review Board gave ethical approval for the Barrett dataset.

### GWAS collection

We downloaded pre-processed summary statistics for the four asthma-associated traits; adult-onset asthma, childhood-onset asthma, all asthma, allergy/eczema and for those in our control panel; height, Alzheimer’s disease and rheumatoid arthritis (**Supplementary Table 1**).

### Summary statistics processing for visualization

We downloaded the childhood-onset asthma and adult-onset asthma ^38^ summary statistics from the GWAS catalog. We used the harmonized summary statistics in the GRCh38 version of the genome. We removed the MHC region (chr6:28510120-33480577).

### Bulk RNA-seq data processing and quality check

FASTQ files were aligned to the GRCh38 or GRCh37 human genome using STAR (v2.7.9a) with standard parameters and two-pass mode, or the Salmon tool (v1.5.1). For BAM files generated with STAR, counts were calculated using RSEM (v1.3.3). We normalized the counts by transforming them to their log2(TPM+1) value, where TPM stands for transcripts per million. To detect outlier samples, we performed principal component analysis on scaled normalized expression for the top 1000 most variable genes that were expressed in at least 25% of the samples. For alignment and quantification, we used the ENSEMBL reference annotation release 105 which was downloaded from the ENSEMBL website.

### Differential expression analyses

Differential gene expression was tested using a linear mixed model, similar to what we did in Gutierrez-Arcelus et al.^69^. Specifically, we used a likelihood ratio test between two nested models (*anova* function in R). In these models, gene expression levels (log2(TPM + 1)) represent the dependent variable. “Donor ID” was included as a predictor variable, treated as a random effect. To compare one condition against the others, we indicated with 1 the tested condition and 0 for the others (the test variable). We used the function “lmer” from the R package “lme4” to implement the model. For risk gene visualization in the miami plot, P-values were corrected for multiple hypothesis testing using the package “qvalue”. Differentially expressed genes at 5% FDR are reported for depicting specific genes in risk loci. After each analysis, we calculated a t-statistic for each gene to rank them and chose the top 10% as annotations for heritability enrichment analysis (see LDSC-SEG section below). The details of each analysis are divided by dataset and described in the following section.

#### - Helling Dataset

To assess DE genes between rhinovirus treatment and PBS vehicle control within healthy individuals, we tested genes that had a normalized count greater than 1 in at least 9 samples which led to a total of 14,883 genes. The threshold for the minimum number of samples reflected half of the biological replicates (9/18). In addition to having “donor ID” as a random effect, we accounted for “sex” as a fixed effect. We repeated this process in asthma patients only, testing 14,888 genes for differential expression between rhinovirus infection and PBS vehicle control. To find DE genes from asthma patients compared to healthy controls, we took all the samples and recalculated the number of genes present in at least 9 samples. We tested 15,935 genes and incorporated “treatment” with either PBS or rhinovirus as a fixed effect covariate in the model, and tested for disease status (0/1).

##### Tao Dataset

We included 16,031 genes having a normalized count greater than 1 in half of the samples (3/6 samples). We tested for differential expression between influenza treatment and control (sham).

##### Koh Dataset

We tested 14,988 genes, to find differentially expressed genes specific to each condition compared to all others: IFNα, IFNγ, IL-13, and IL-17, respectively. We selected genes having a normalized count greater than 1 in at least 6 samples. This number reflected the smallest amount of replicates found across conditions (6/36 samples).

#### - Basnet Dataset

We excluded the resting sample from donor B03 from these analyses (see Methods, Bulk RNA-seq, QC, and analysis). To obtain differentially expressed gene profiles for each time point, we performed four separate models in which we tested a single time point against the other two and the resting condition. Our fourth model tested for DE genes in activation conditions versus the resting state through the same approach. Since we had 3 biological replicates for most time points, we tested genes with a normalized count greater than 1 in at least (3/11) samples, which yielded 14,380 genes. This gene set was used for all models.

#### - Barrett Dataset

Air-liquid interface (ALI) cultures were grown from nasal basal epithelial cells from 6 healthy adult donors. ALIs were allowed to mature for 14 days, then stimulated with 10 ng/mL of IL-4 and 10 ng/mL of IL-13 for an additional 7 days, and then lysed with TCL buffer (Qiagen 1031576) at the conclusion of the experiment. Lysates were stored at −80C and later submitted to the Broad for SmartSeq2 low input bulk RNA-seq (38bp paired-end sequencing).

To obtain genes DE under IL-4 and IL-13 co-stimulation we compared them against the non-stimulated cells. We tested 13,518 genes that had a normalized count greater than 1 in at least half of the samples (9/18) samples.

### ATAC-seq data processing and differential accessibility analysis

We used the 829,942 consensus peaks called by Calderon et al. (peaks were called in each sample separately, then merged across samples, and then counts were re-calculated for all samples using the merged peak coordinates). We transformed counts into reads per kilobase per million (RPKM), then quantile normalized and finally scaled to their log2(normalized RPKM+1). To assess differentially accessible (DA) peaks, we first calculated the mean normalized count per cell type and then created cell-type accessible sets of peaks. We included a peak in the set if it had a normalized count greater than the mean of the cell type in at least half of the samples corresponding to subtypes of that cell type. We tested between 400 and 600 thousand peaks per cell-type for DA. To do so, we implemented a linear mixed model using the normalized counts as the response variable, and for the predictor, a bit flag system indicating 1 if the sample belonged to the tested cell type and 0 for the remaining cells. Peaks were sorted by t-statistic and we took the top 10% peaks for each cell-type-specific annotation. This process was replicated for a second selection model, implemented to divide peaks between stimulated and unstimulated categories.

### Single-cell RNA-seq, QC and analysis

FASTQ files from the single-cell RNA-seq dataset from Wang et al., Seumois et al., Basnet et al., and Ravindra et al., ^35,70–72^ were downloaded as indicated in the “Dataset Collection” section. For each dataset, we aligned FASTQ files to the human reference human genome GRCh38 ^73^, using the GENCODE release 32 with cellranger count (v6.1.2), using default parameters. For Basnet et al, we added the RV-C15 sequence to the reference genome. Counts were then aggregated using the cellranger aggr (v6.1.2) function with the default parameters. The subsequent analyses were done for each of the datasets individually. We discarded cells with less than 500 genes expressed and cells expressing more than 20% of mitochondrial genes. We normalized raw counts with the “LogNormalize” method from Seurat package (v4.0.5). We used the normalized counts to perform a PCA with the 1000 most variable genes. We used the top 20 principal components to perform dimensionality reduction with UMAP to visualize the data. We identified clusters using the “FindClusters” function with the Louvain algorithm and a resolution parameter of 0.2, using the top 20 principal components (PCs). We corrected PCs with Harmony package ^74^ (v0.1.1) as indicated as follows, if nothing is indicated, no corrections were applied. For Wang et al., dataset we corrected for “donor ID” and “tissue”. For Basnet et al., dataset we corrected for “donor ID” and virus infection. For Ravindra et al., dataset we corrected for virus infection.

To identify which cellular type was present in each cell cluster, we used the function “FindVariableFeatures” (parameter; test.use=wilcox) from Seurat package to identify differentially expressed genes in each cluster. If immune cells were present in the dataset we used the tool MCPcounter (v1.2.0) to annotate immune cells ^75^. Based on these cellular markers we annotated the clusters with data from the literature ^35,76,77^.

For data from Ziegler et al.^40^ we used the UMAP coordinates, and the cell annotation originally published by the authors.

### LDSC-SEG

State-specific gene sets were generated using the top 10% of the genes tested ranked by t-statistic for each of our DE analyses. Genes coordinates were mapped from the human genome reference GRCh37 GTF file and formatted into bed files. We repeated this process to generate control bed files containing all the genes tested for DE in each analysis. Symmetric windows of 100kb were added at each side of the genes using the bedtools (v2.31) “slop” function. For bed files containing ATAC-seq peaks, this window consisted of 225 bp at each side of the peak, to represent a similar genomic coverage. LD-Score files were generated using the LDSC pipeline along with data from HapMap 3 and Phase 3 of the European 1000 Genomes obtained from the LDSC-repository. The regression was run using the baseline model v1.2. We reported the P values of regression coefficients, and normalized regression coefficient as per-standardized-annotation effect sizes τ* as in (Gazal et al. 2017 Nat Genet)^78^ to allow for multi-trait comparisons. Regression P values were corrected using the “p.adjust” function from R using both FDR and bonferroni methods. The reference GTF file used to map the genes was obtained from GENCODE (v37).

### MAGMA and scDRS

We used the adult-onset asthma, the childhood-onset asthma, the allergy/eczema, the all-asthma, the rheumatoid arthritis, and the Alzheimer’s GWAS summary statistics as well as the corresponding set of 1000 putative disease genes (obtained with MAGMA) provided in the original publication of the scDRS method ^17,79^.

We used MAGMA (v1.10) to compute the gene-level association P-values and z-scores from GWAS summary statistics of Height. We transformed P-values to z-scores using this formula: 2*pnorm(abs(zscore), mean = 0, sd = 1, lower.tail = F) in R. To map SNPs to genes, we used magma with default parameters specified in the scDRS documentation. We retrieved the top 1000 genes based on MAGMA z-score as putative disease genes.

We used scDRS (v1.0.2) to quantify the expression of the putative disease genes derived from GWAS summary statistics using MAGMA in each cell of each single-cell RNA-seq dataset separately for the 8 GWAS tested and described previously (Table dataset). We used the function scDRS “compute-score” with default parameters (--flag-filter-data False).

### Cell-cell interaction between ciliated epithelial cells and non-ciliated epithelial cells

To identify potential pairs of interactors between ciliated and non-ciliated epithelial cells we used the dataset of Basnet et al. ^35^. We first evaluated which genes were differentially expressed between rhinovirus infected epithelial cells and non-ciliated non-infected epithelial cells. We identified the hypothetical pair of interactors (ligand-receptor) between ciliated and non-ciliated cells with CellPhoneDB (v3.1.0) with the method “deg_analysis”, using the list of differentially expressed genes defined before and the default parameter. We filtered the results by identifying the ligand being expressed in ciliated cells and a receptor expressed in non-ciliated cells.

### Linking variants to genes

For the Miami plot we used three different and complementary approaches to map variants to genes. First, we used data from Open Target Genetics website ^80^. We queried the website to retrieve information for childhood-onset asthma (GCST007995) and adult-onset asthma (GCST007799). We then extracted the L2G gene and the Closest Gene for each variant when information was available. We also added a window of 250kb around each variant with the “bedtools slop” function and retrieved the genes falling in those regions with the “bedtools intersect” function. Finally, we obtained the intersect between this “snp-to-gene” list of genes and the genes upregulated upon rhinovirus infection (genes annotated on **Figure 4A**).

We also retrieved the lead variants from the original paper from Ferreira et al. ^38^. We added a window of 250kb around each variant with the “bedtools slop” function and retrieved the genes falling in those regions with the “bedtools intersect” function. We identified the closest gene to the variant by using the function “bedclosest”.

### Software description (Plots, R, and Biorender)

All the plots were generated with R and graphic schematics were generated with Biorender.

### Data availability

Bulk RNA-seq data on dataset with IL-4 and IL-13 activation of airway epithelial cells will be made available in GEO and dbGAP. Other datasets used are publicly available (details in **Supplementary Table 1**).

## Data Availability

All data produced in the present work are contained in the manuscript and are available upon reasonable request to the author

## Acknowledgements

This study was supported by U19AI095219. CO was supported by U19AI162310. NAB was supported by AI134989 and U19AI095219. JG and NAB were supported by AADCRC RNA Sequencing Core for Airway Epithelial Cells. MGA was supported by P30AR070253. We thank Donata Vercelli, Luis Barrera, Peter Nigrovic and his laboratory, and the Gutierrez-Arcelus laboratory for feedback on this study.

## Graphical abstract

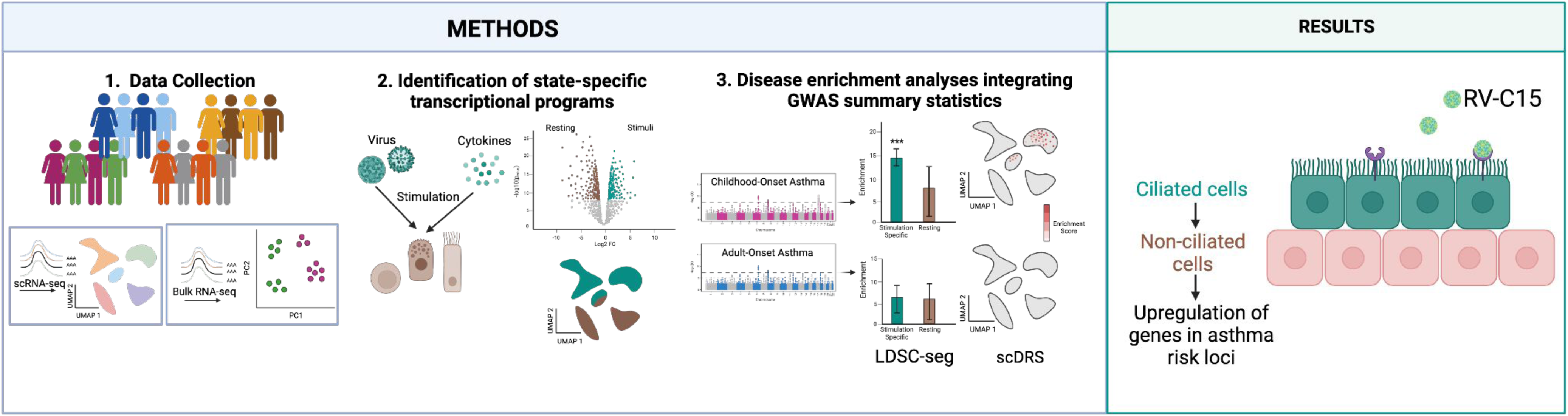

---

**Supplementary Figure 1.**
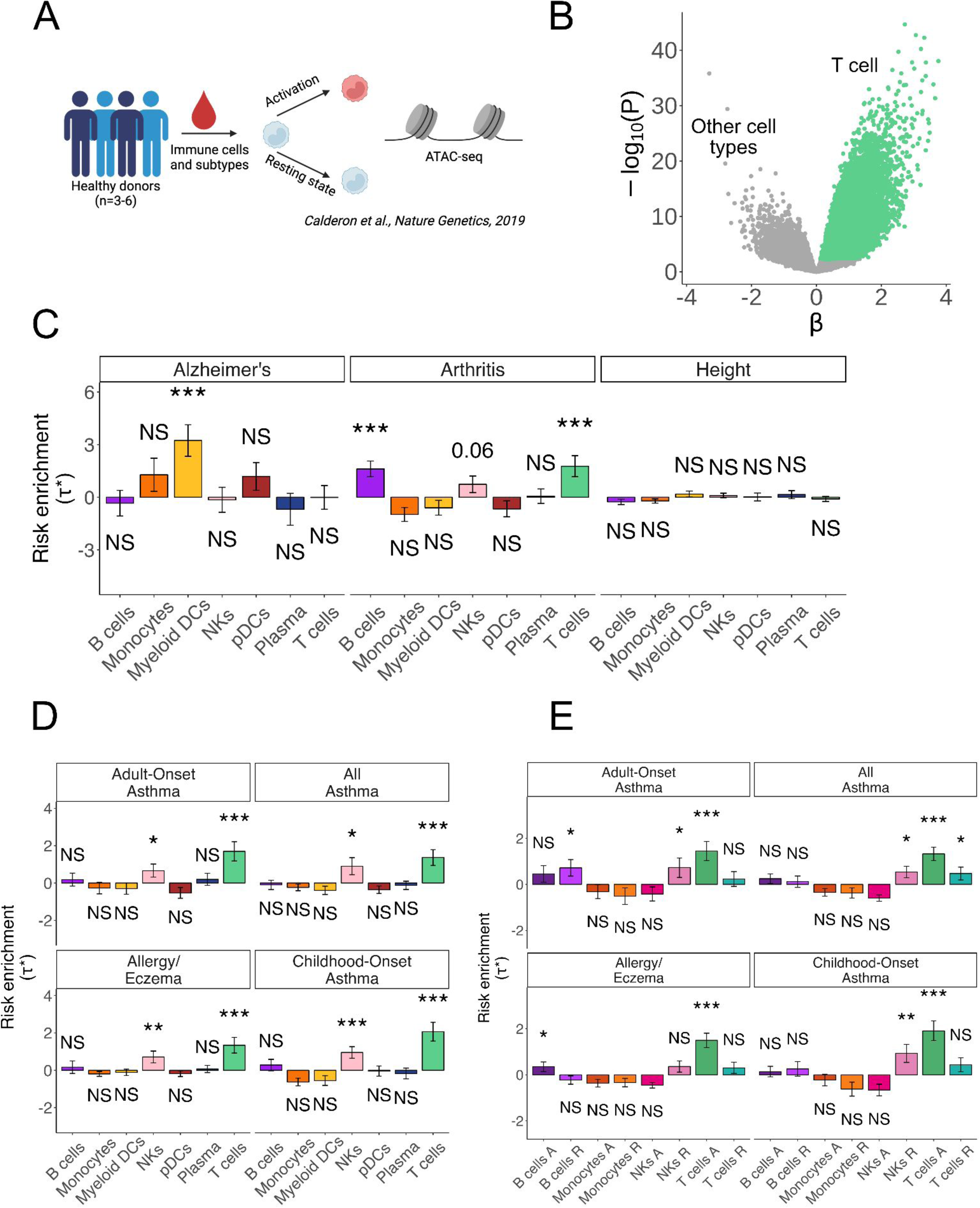
Validation of T cells enrichment in asthma-associated loci using differentially accessible peaks. **(A)** Experimental design of the *Calderon et al.* ATAC-seq dataset of immune cell types that were activating or not *in vitro*. **(B)** Volcano plot showing differentially expressed peaks between T cells and all other cell types. Peaks upregulated in T cells were selected based on t-statistic and are colored in green. **(C)** Bar plots representing LDSC-SEG heritability enrichment coefficient (τ*) for each cell type for the 3 control traits tested. **(D)** Bar plot representing LDSC-SEG heritability enrichment coefficient (τ*) for each set of cell-type-specific differentially accessible peaks for each of the asthma-associated traits. **(E)** Bar plots showing LDSC-SEG heritability enrichment coefficient (τ*) for each of the asthma-associated traits in cell-state-specific DA peaks of immune cells divided in either resting (light colors) or activated (dark colors) condition. In all bar plots, error bars represent τ* +/-standard error and asterisks denote significance as *** Bonferroni-adjusted P < 0.05, ** FDR 5%, * P < 0.05, and NS denotes nonsignificant (P > 0.05).

**Supplementary Figure 2.**
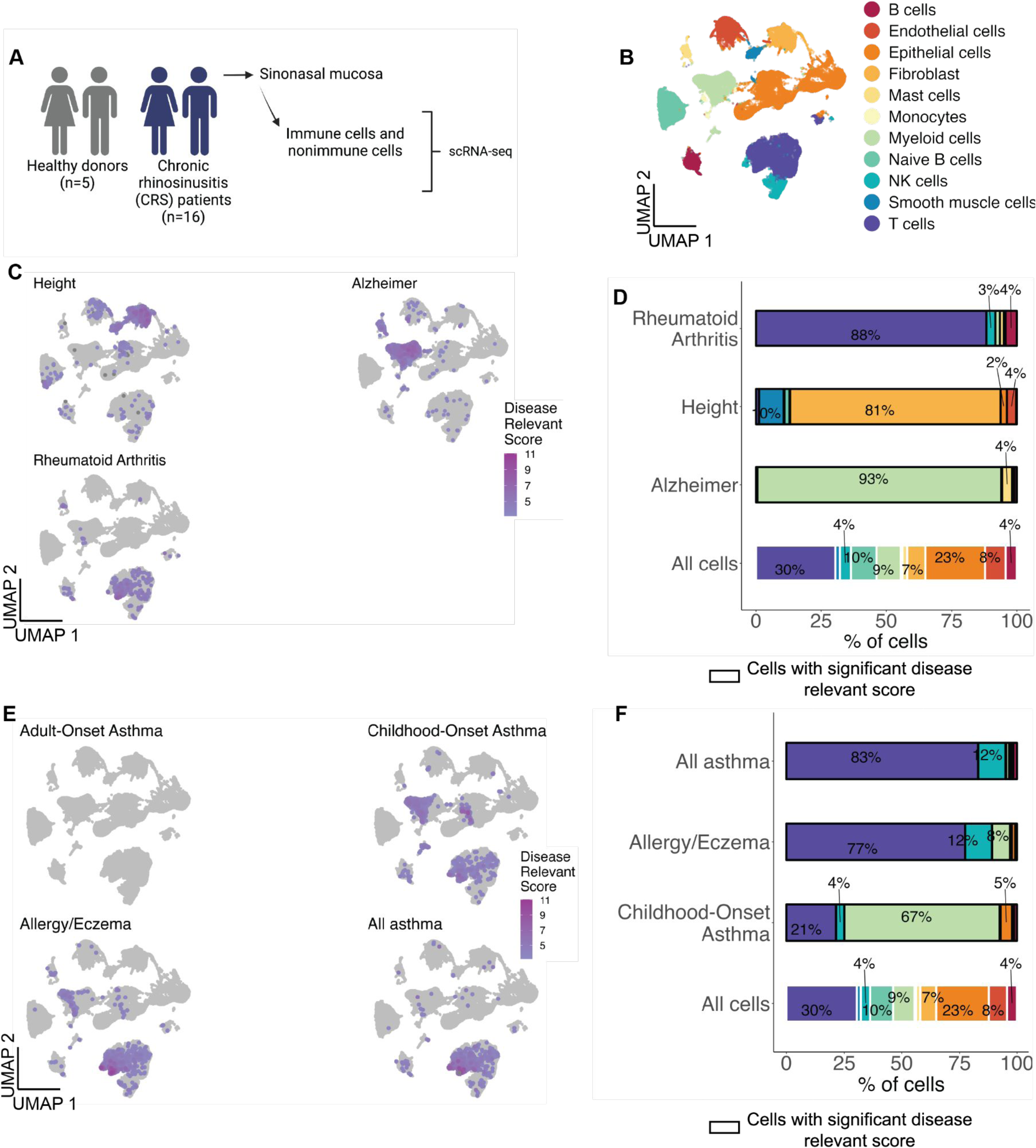
T cell validation at the single-cell RNA-seq level. **(A)** Experimental design of the *Wang et al.* scRNA-seq dataset of sinonasal mucosa from healthy donors and chronic rhinosinusitis patients (CRS). **(B)** UMAP visualization of the 1,115,856 immune and non-immune cells colored by cell type. **(C)** scDRS results represented on the UMAP for the 3 control GWAS tested. The intensity of the color represents the disease relevant score, the lighter purple represents a less intense score whereas a more intense purple represents cells associated with a stronger score. Non-significant cells with a FDR higher than 10% are depicted in gray. **(D)** Bar plot representing the percentage of each cell type in all cells followed by the significant cells at 10% FDR for scDRS in Alzheimer’s Disease, Height and Rheumatoid Arthritis. **(E)** scDRS results represented on the UMAP for the 4 asthma-associated traits tested. The intensity of the color represents the disease relevant score, the lighter color represents a less intense score whereas a more intense color represents cells associated with a stronger score. **(F)** Bar plot representing the percentage of each cell type in all cells followed by the significant cells at 10% FDR for scDRS in COA, Allergy/Eczema and All asthma.

**Supplementary Figure 3.**
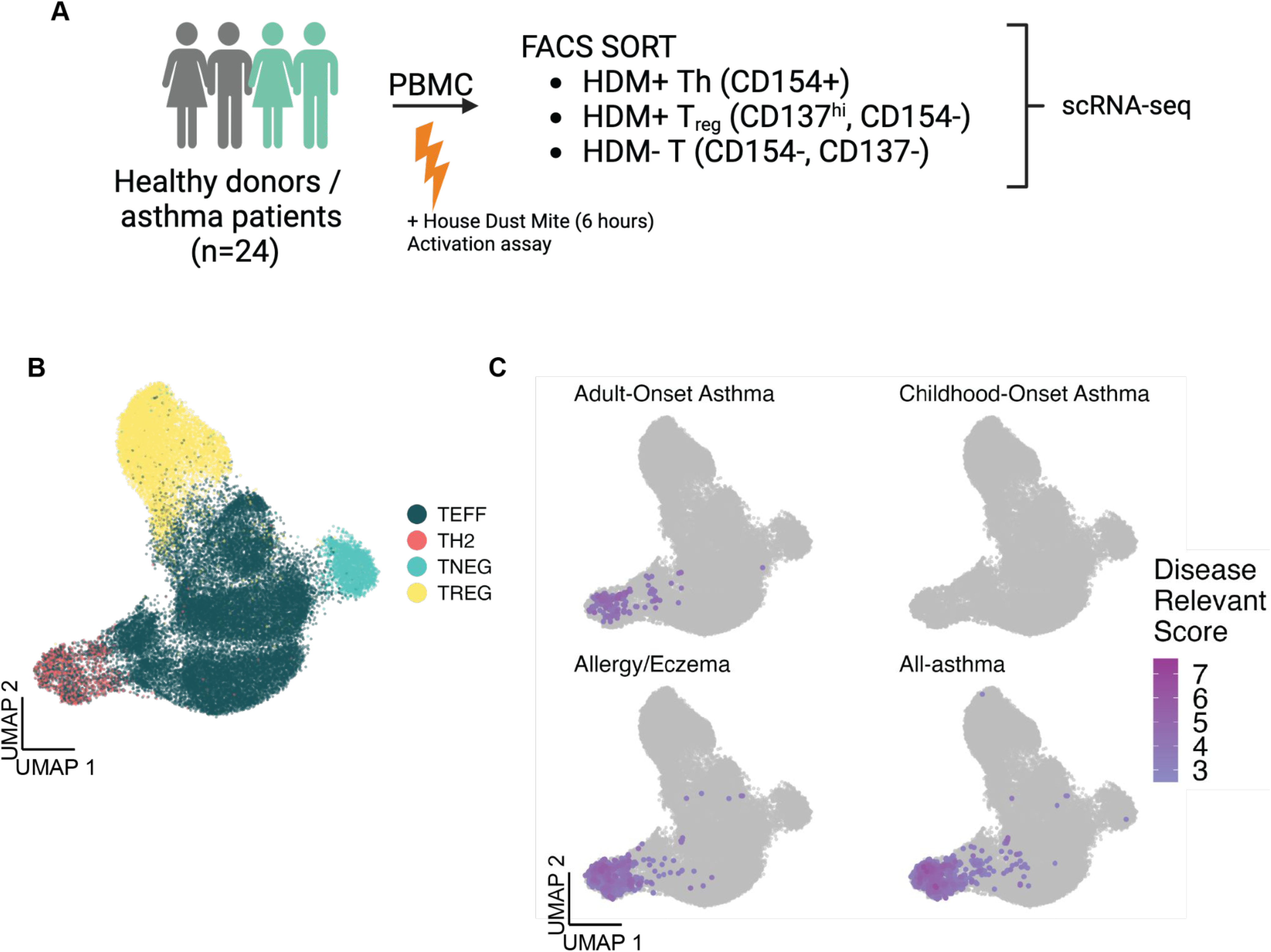
Th2 validation at the single-cell RNA-seq level. **(A)** Experimental design of the *Seumois et al.* scRNA-seq dataset consisting of T cells. **(B)** UMAP visualization of the 38,559 T cells colored by subtypes. **(C)** scDRS results represented on the UMAP for the 4 asthma-associated GWAS tested. The intensity of the color represents the disease relevant score, the lighter color represents a less intense score whereas a more intense color represents cells associated with a stronger score. nonsignificant cells with a FDR higher than 10% are depicted in gray.

**Supplementary Figure 4.**
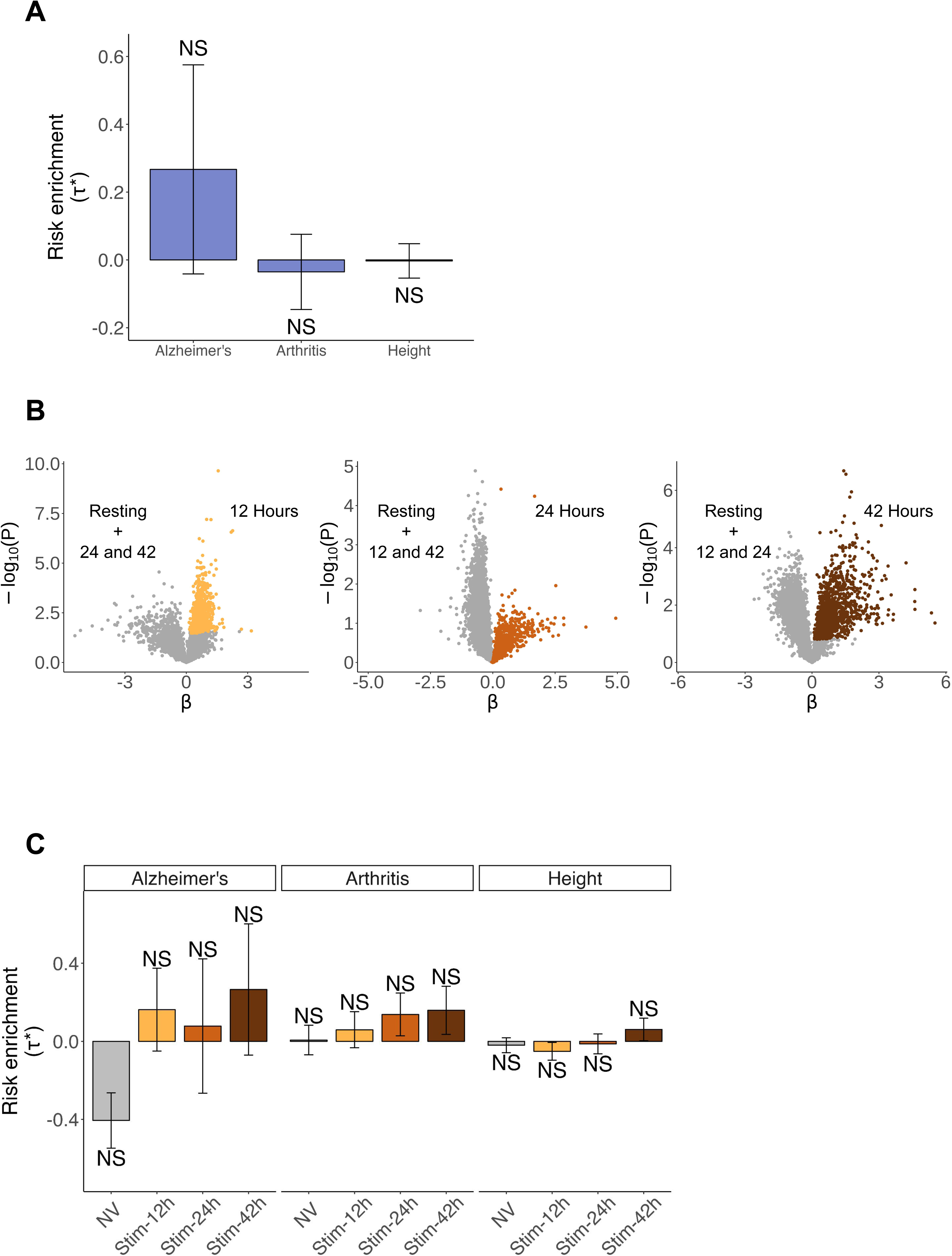
Control traits of BECs from healthy individuals infected with RV. **(A)** Bar plots representing LDSC-SEG heritability enrichment coefficient (τ*) for the 3 control traits. NS denotes nonsignificant (P > 0.05). **(B)** Volcano plots showing differentially expressed genes at each time point (12,24,42) compared to all others in RV-infected epithelial cells colored in yellow, orange and brown respectively. **(C)** Bar plot representing LDSC-SEG heritability enrichment coefficient (τ*) for each time point against all others for each of the control trait tested. NS denotes nonsignificant (P > 0.05). In all bar plots, error bars represent τ* +/-standard error.

**Supplementary Figure 5.**
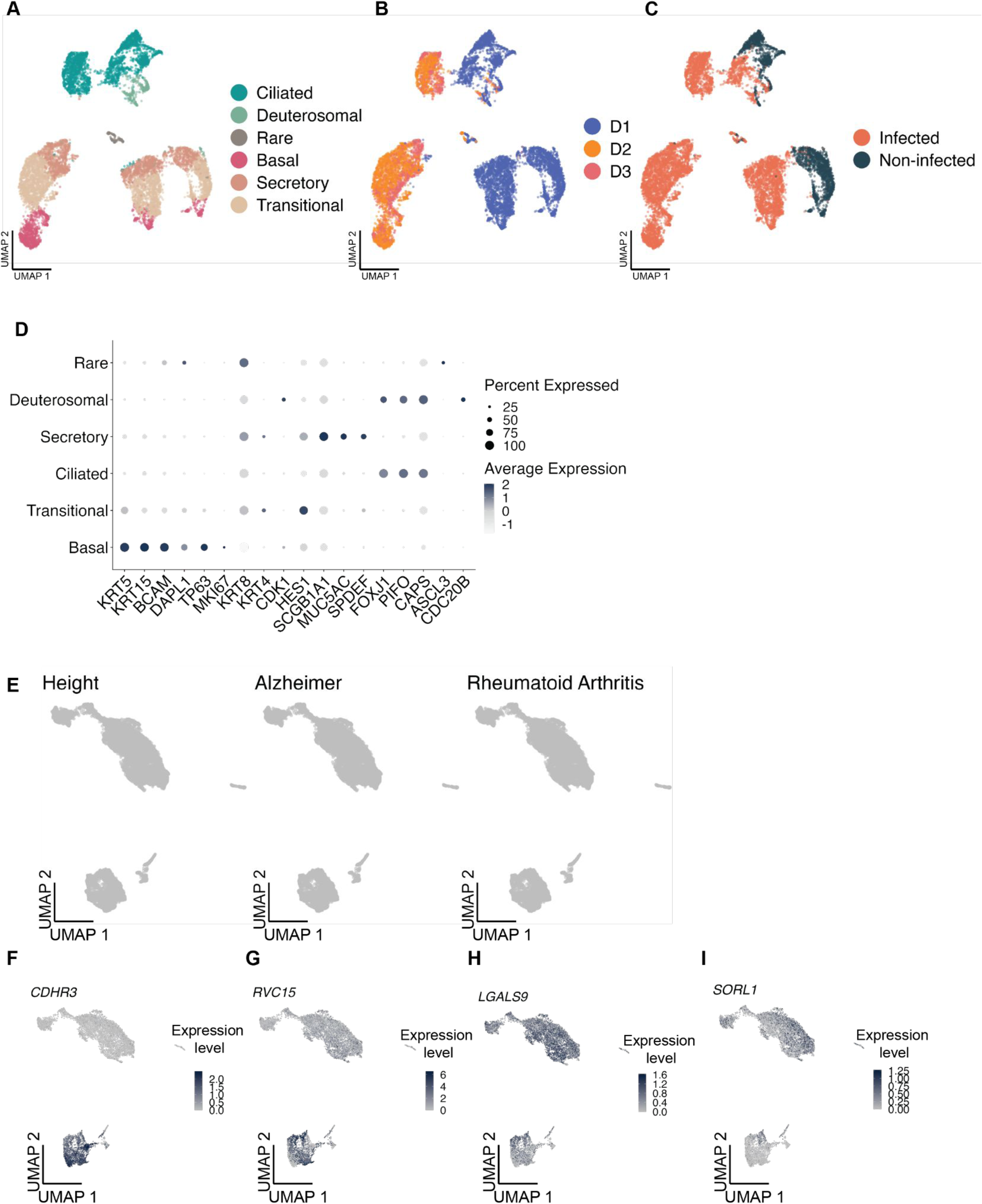
Additional informations related to Figure 2. UMAP visualization of the 10,721 epithelial cells colored by **(A)** cell type. **(B)** donor. **(C)** infection status (Infected / Non-infected). **(D)** Dot plot representing the normalized average expression and the percent of cells expressing a given gene for epithelial cell markers. **(E)** scDRS results represented on the UMAP for the 3 control GWAS tested. nonsignificant cells with a FDR higher than 10% are depicted in gray. **(F-I)** Harmonized UMAP representing *CDHR3, RVC15, LAGLS9 and SORL1* normalized expressions.

**Supplementary Figure 6.**
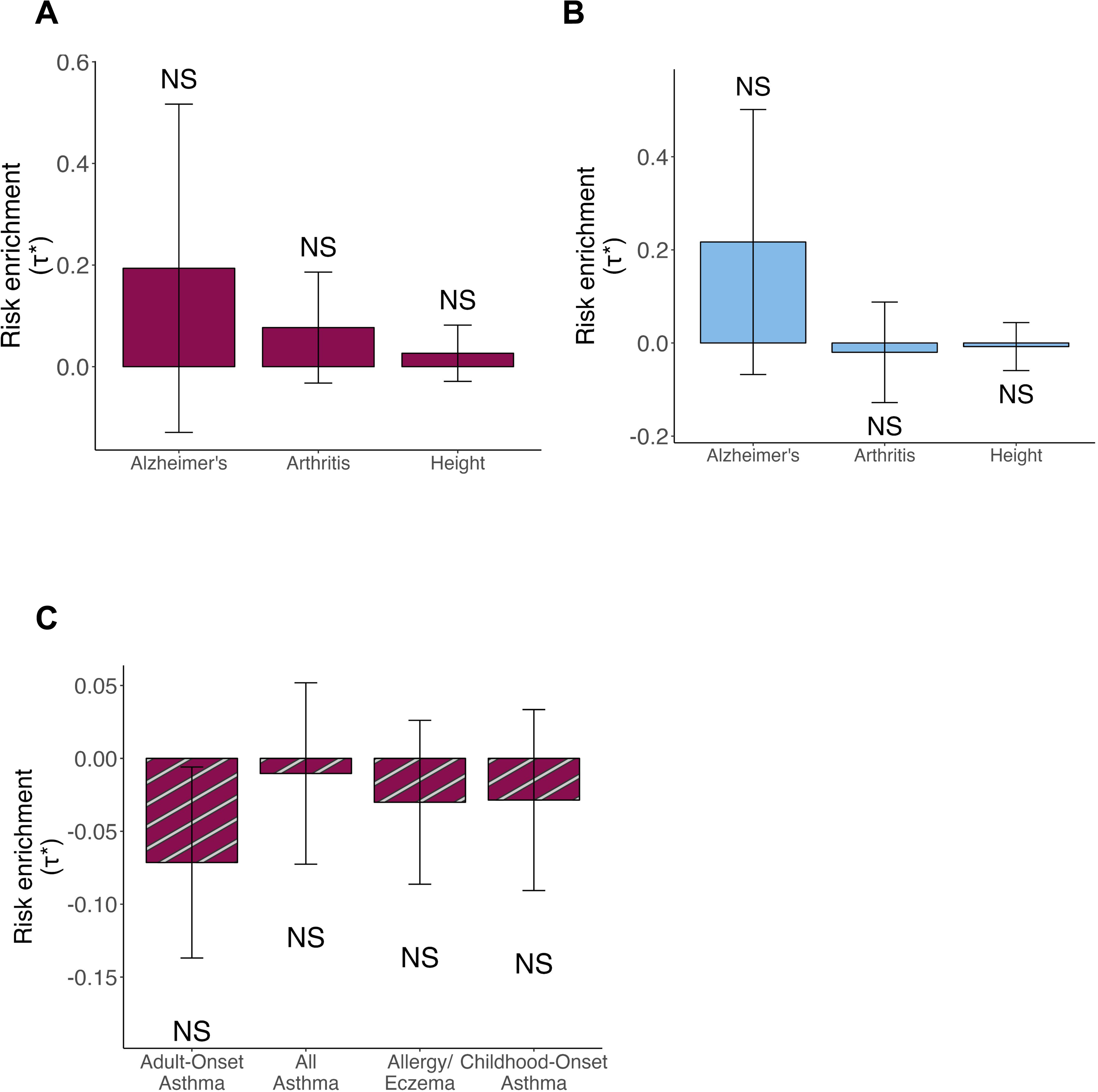
Control traits of BECs from asthma patients infected with RV. Bar plots representing LDSC-SEG heritability enrichment coefficient (τ*) for the 3 control traits tested. NS denotes nonsignificant (P > 0.05). **(A)** Using DE genes after RV-infection in epithelial cells from patients. **(B)** Using genes differentially expressed in asthmatics when compared to healthy individuals. **(C)** Bar plot showing LDSC-SEG heritability enrichment coefficient (τ*) across traits for genes downregulated upon RVC-15 infection. NS denotes nonsignificant (P > 0.05). In all bar plots, error bars represent τ* +/-standard error.

**Supplementary Figure 7.**
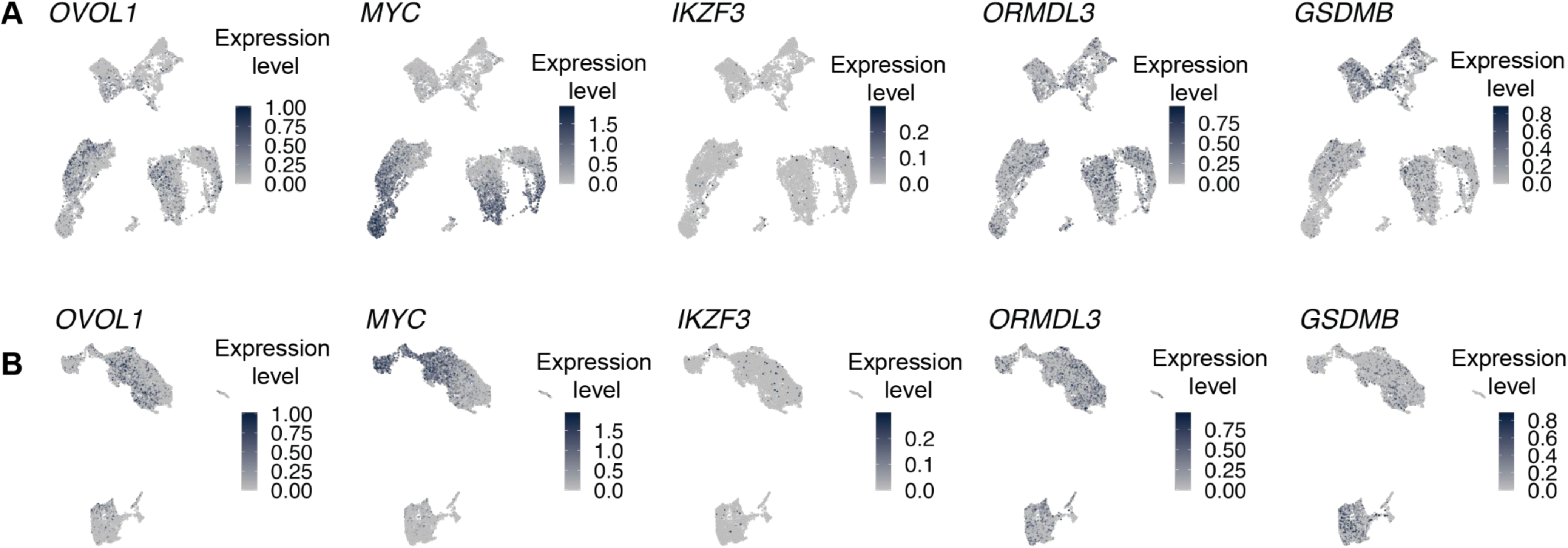
Normalized expressions of gene of interest. **(A)** Normalized expression of genes of interest represented on the UMAP and **(B)** on the harmonized UMAP.

**Supplementary Figure 8.**
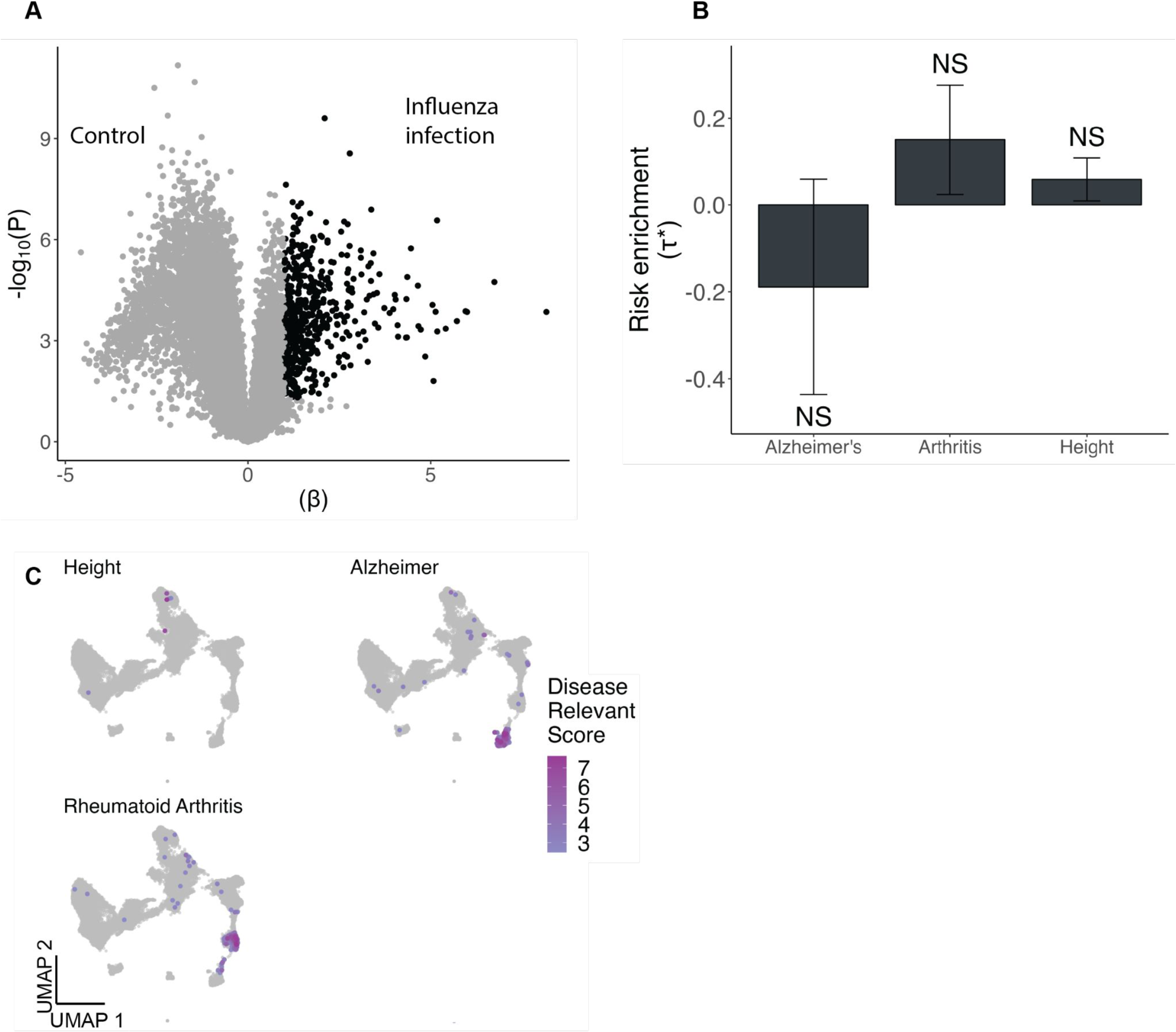
Volcano plot and controls traits related to Figure 5. **(A)** Volcano plot showing differentially expressed genes between influenza infection and control. Genes upregulated upon influenza infection were selected based on t-statistic and are colored in black. **(B)** Bar plots representing LDSC-SEG heritability enrichment coefficient (τ*) for the 3 control GWAS tested. Error bars represent τ* +/-standard error. NS denotes nonsignificant (P > 0.05). **(C)** scDRS results represented on the UMAP for the 3 control traits tested. The intensity of the color represents the disease relevant score, the lighter color represents a less intense score whereas a more intense color represents cells associated with a stronger score. nonsignificant cells with a FDR higher than 10% are depicted in gray.

**Supplementary Figure 9.**
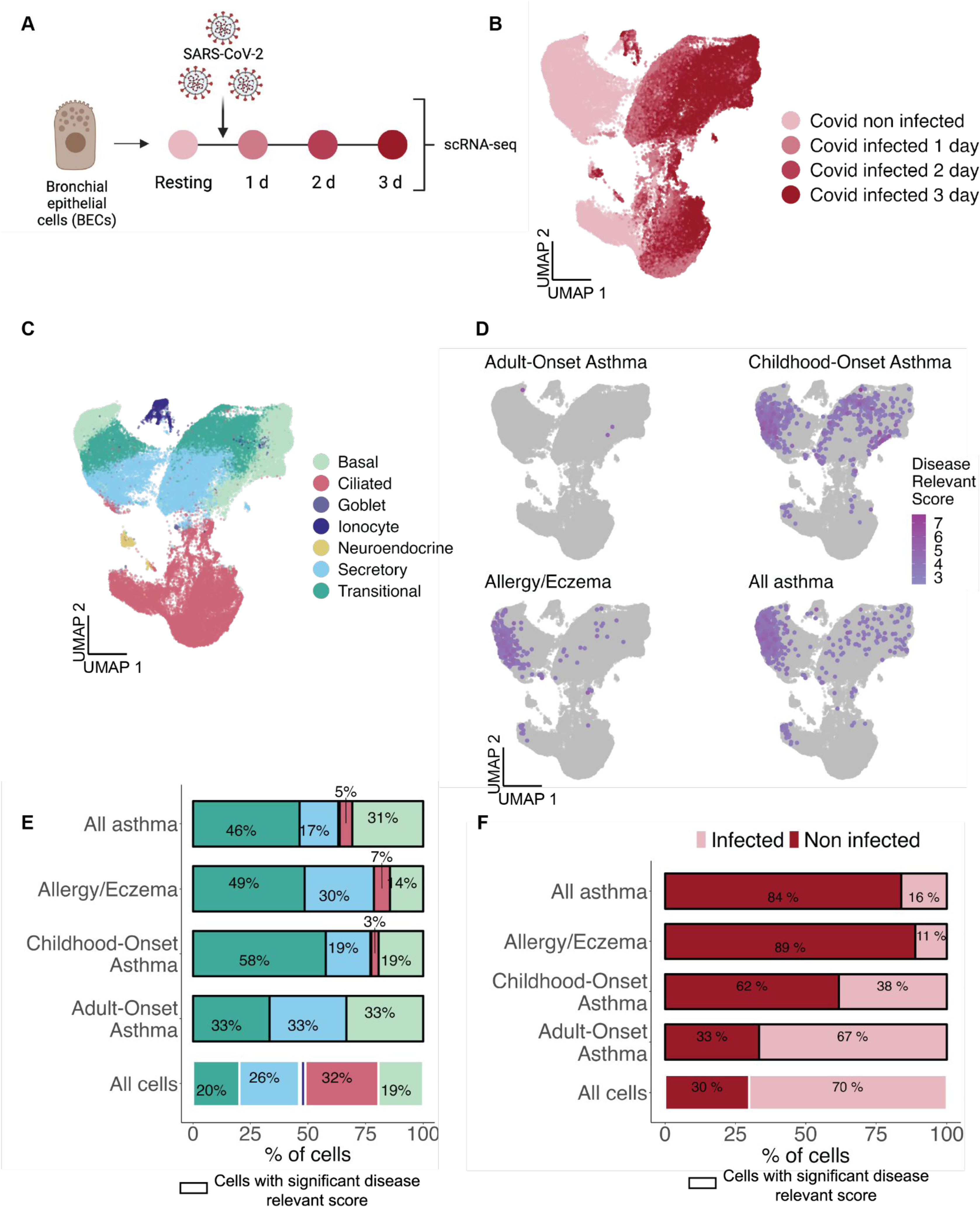
scRNA-seq analysis of BECs infected or not with SARS-CoV-2. **(A)** Experimental design of the *Ravindra et al.* scRNA-seq dataset of bronchial epithelial cells infected with SARS-CoV-2 or not, from one healthy donor. UMAP visualization of the 74,088 cells colored by **(B)** days after virus infection and **(C)** by cell type. **(D)** scDRS results represented on the UMAP for the 3 control traits tested. The intensity of the color represents the disease relevant score, the lighter color represents a less intense score whereas a more intense color represents cells associated with a stronger score. nonsignificant cells with a FDR higher than 10% are depicted in gray. **(E)** Bar plot representing the percentage of each cell type in all cells followed by the significant cells at 10% FDR for scDRS in AOA, COA, Allergy/Eczema and All Asthma. **(F)** Bar plot representing the percentage of cells in the full dataset classified in SARS-CoV-2 infection or non infected, followed by percentage of cells passing significance at 10% FDR for AOA, COA, Allergy/Eczema, All asthma, by SARS-CoV-2 infection or not.

**Supplementary Figure 10.**
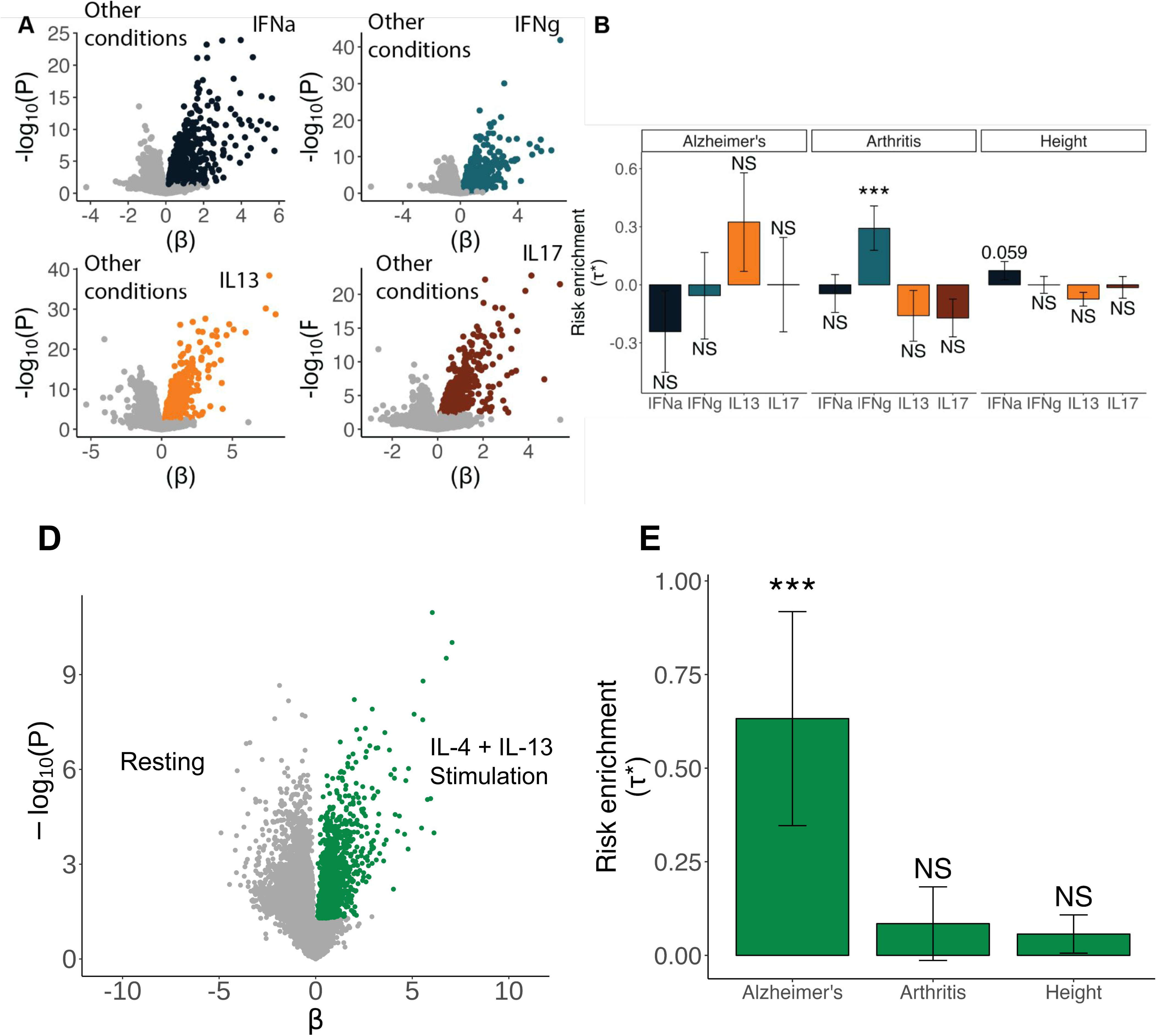
Control traits for Figure 6. **(A)** Volcano plots showing differentially expressed genes for each stimuli (IFNα, IFNγ, IL-13 and IL-17) compared to all others in bronchial epithelial cells colored in black, teal, orange and brown respectively. **(B)** Bar plot representing LDSC-SEG enrichment for each stimuli against all others for each of the 3 control traits. Error bars represent τ* +/-standard error. Asterisk denotes significance as * P < 0.05 and NS denotes nonsignificant (P > 0.05). **(D)** Volcano plot showing differentially expressed genes between IL-4/IL-13 stimulation and resting condition. Genes upregulated upon IL-4/IL-13 stimulation were selected based on t-statistic and are colored in green. **(E)** Bar plots representing LDSC-SEG heritability enrichment coefficient (τ*) for the 3 control traits. Error bars represent τ* +/-standard error. Asterisk denotes significance as * P < 0.05 and NS denotes nonsignificant (P > 0.05).

